# Improved Detection of Decreased Glucose Handling Capacities via Novel Continuous Glucose Monitoring-Derived Indices: AC_Mean and AC_Var

**DOI:** 10.1101/2023.09.18.23295711

**Authors:** Hikaru Sugimoto, Ken-ichi Hironaka, Tomoaki Nakamura, Tomoko Yamada, Hiroshi Miura, Natsu Otowa-Suematsu, Masashi Fujii, Yushi Hirota, Kazuhiko Sakaguchi, Wataru Ogawa, Shinya Kuroda

**Affiliations:** Department of Biochemistry and Molecular Biology, Graduate School of Medicine, The University of Tokyo, 7-3-1 Hongo, Bunkyo-ku, Tokyo 113-0033, Japan; Department of Biological Sciences, Graduate School of Science, The University of Tokyo, 7-3-1 Hongo, Bunkyo-ku, Tokyo 113-0033, Japan; Department of Diabetes and Endocrinology, Akashi Medical Center, 743-33 Okubo-cho Yagi, Akashi, Hyogo 674-0063, Japan; Division of Diabetes and Endocrinology, Department of Internal Medicine, Kobe University Graduate School of Medicine, 7-5-1 Kusunoki-cho, Chuo-ku, Kobe, Hyogo 650-0017, Japan; Department of Diabetes and Endocrinology, Takatsuki General Hospital, 1-3-13 Kosobe-cho, Takatsuki, Osaka 569-1192, Japan; Department of Mathematical and Life Sciences, Graduate School of Integrated Sciences for Life, Hiroshima University, 1-3-1 Kagamiyama, Higashi-hiroshima City, Hiroshima 739-8526, Japan

**Keywords:** Continuous glucose monitoring, oral glucose tolerance test, clamp test, disposition index, insulin clearance

## Abstract

**Background:** Efficiently detecting impaired glucose control abilities is a pivotal challenge in public health. This study assessed the utility of relatively easy-to-measure continuous glucose monitoring (CGM)-derived indices in estimating glucose handling capacities calculated from resource-intensive clamp tests.

**Methods:** We conducted a prospective, single-center, observational cohort study of 64 individuals without prior diabetes diagnosis. The study utilized CGM, oral glucose tolerance test, and hyperglycemic and hyperinsulinemic-euglycemic clamp tests. We validated CGM-derived indices characteristics using an independent dataset and mathematical model with simulated data.

**Findings:** A novel CGM-derived index, AC_Var, was significantly correlated with insulin sensitivity (r = –0.31; 95% CI: –0.52 to –0.06), insulin clearance (r = –0.31; 95% CI: –0.54 to –0.06), and disposition index (DI) (r = –0.31; 95% CI: –0.52 to –0.07) calculated from clamp tests.

AC_Var was also significantly correlated with insulin resistance (r = 0.48; 95% CI: 0.23 to 0.68) in an independent dataset. Multivariate analyses indicated AC_Var’s contribution to predicting reduced blood glucose control abilities independent from conventional CGM-derived indices. The prediction model’s accuracy utilizing CGM-measured glucose standard deviation and AC_Var as input variables, with clamp-derived DI as the outcome, closely matched that of predicting clamp- from OGTT-derived DI. Mathematical simulations also underscored AC_Var’s association with insulin clearance and DI.

**Interpretation:** CGM-derived indices, including AC_Var, can be useful for screening decreased blood glucose control ability. We developed a web application that calculates these indices (https://cgm-ac-mean-std.streamlit.app/).

**Funding:** The Japan Society for the Promotion of Science KAKENHI, CREST, Japan Science and Technology Agency, and Uehara Memorial Foundation.

## INTRODUCTION

Pre-diabetes (pre-DM), a condition in which blood glucose controlling mechanisms are slightly disrupted, is a high-risk condition for the development of type 2 diabetes mellitus (T2DM) and its associated complications.^1^ Approximately 5–10% of patients with pre-DM develop T2DM annually,^1,2^ with up to 70% eventually developing T2DM.^1,3^ Despite lower blood glucose levels in pre-DM compared to T2DM, individuals with pre-DM are at elevated risk of chronic kidney disease^4^ and cardiovascular disease,^5^ which are common complications of diabetes. However, preventive measures through pharmaceutical or lifestyle interventions can impede the progression to T2DM and mitigate complications.^1,6,7^ Consequently, pre-DM screening followed by targeted interventions has demonstrated cost-effectiveness.^8^

Optimal methods for detecting the decreased ability to control blood glucose, however, have yet to be established. Hemoglobin A1c (HbA1c) and fasting blood glucose (FBG) have been recommended for screening for pre-DM, but they lack precision for prediabetes screening and even their combination has a diagnostic sensitivity of only about 60%.^9–11^ The 2-h oral glucose tolerance test (OGTT) has long been the gold standard diagnostic method for pre-DM and T2DM.^12^ However, the cost, time, patient inconvenience, and lack of reproducibility of the OGTT make it impractical for patient care.^12^ Moreover, these indices lack information on glycemic variability and changes in glucose levels throughout the day, which are relevant to T2DM complications.^13,14^ Insulin sensitivity and insulin secretion impairment precede T2DM development, with insulin resistance emerging years prior and decreased β-cell function already present in pre-DM.^1^ Although hyperinsulinemic-euglycemic and hyperglycemic clamp tests have been the gold standards for assessing insulin sensitivity and insulin secretion,^15^ their resource intensiveness impedes practicality in screening. Consequently, the development of more accurate and accessible methods for detecting reduced glucose control abilities constitutes a pressing public health challenge.

Continuous glucose monitoring (CGM) has emerged as a promising avenue for identifying individuals with reduced glucose handling capacity.^16–18^ CGM-derived indices identify individuals who are not identified as pre-DM by FBG, HbA1c, or OGTT, but who have high levels of postprandial glucose, a characteristic of pre-DM and even T2DM.^19^ A CGM-derived index has demonstrated the ability to identify individuals with a high risk of progressing to T2DM.^20^ Some CGM-derived indices also correlate with insulin secretion, insulin sensitivity, or the disposition index (DI).^19,21–24^ While various glycemic variability indices derived from CGM have been reported, their relationship to glucose handling capacity and their potential for identifying individuals with reduced glucose handling, particularly in a population predominantly composed of healthy individuals, remain unclear.

Here, we assessed previously established CGM-derived indices^19,25^ and introduced two novel indices (AC_Mean and AC_Var) based on autocorrelation function of glucose levels. By examining the correlation between these indices and indices calculated from OGTT or clamp tests, we found that AC_Mean and AC_Var were significantly correlated with insulin clearance or the DI, unlike the other CGM-derived indices. In addition, multivariate analyses indicated that CGM-derived indices, including AC_Mean and AC_Var, contributed to the prediction of decreased blood glucose control abilities. A mathematical model with simulated data also indicated that AC_Mean and AC_Var were associated with insulin clearance and the DI.

## METHODS

### Subjects and measurements

This study was conducted in accordance with the Declaration of Helsinki and its amendments, and was approved by the ethics committee of Kobe University Hospital (Approval No. 1834; Kobe, Japan). Written informed consent was obtained from all subjects. Study participants who had no previous diagnosis of diabetes and were over 20 years old were recruited from Kobe University Hospital (Hyogo, Japan) from January 2016 to March 2018. Exclusion criteria were: 1) taking medications that affect glucose metabolism (e.g., steroids, β blockers); 2) patients with psychiatric disorders; 3) pregnant or breast-feeding women; and 4) deemed unfit for any other reason by attending physicians.

The study participants initially underwent a 75-g OGTT in the morning after an overnight fast. Following the OGTT, they wore a CGM device (iPro; Medtronic, Minneapolis, MN, USA) for more than 72 h. Within 7 days after the OGTT, the participants underwent a consecutive hyperglycemic and hyperinsulinemic-euglycemic clamp test. Detailed procedures for the OGTT, consecutive hyperglycemic and hyperinsulinemic-euglycemic clamp, and CGM can be found in Supplementary Text 1.

A total of 70 participants were initially enrolled. One participant taking a β blocker, two participants with missing CGM data, two participants with protocol deviation, and one participant with missing OGTT and/or clamp data were excluded from the analysis.

Consequently, data from 64 participants were used in the analysis (Fig. S1). The sample size of 64 participants closely aligned with the cohort of 57 individuals investigated in a previous study, where statistically significant correlations between CGM-derived indices and the ability to regulate blood glucose were demonstrated.^19^ Of note, with a type I error of 0.05, a power of 0.8, and an expected Spearman correlation coefficient of 0.35, a sample size of 66 (Bonett and Wright’s method) or 64 (Caruso and Cliff’s method) was required to detect a significant difference from zero in the correlation coefficient. This sample size estimation was performed using SPSS version 29 (SPSS Inc.).

### Subjects and measurements of a previously reported dataset

We also performed an analysis using publicity available data sets of CGM (Dexcom G4 CGM System; Dexcom, Fort Lauderdale, FL, USA), OGTT, and steady state plasma glucose (SSPG) test outcomes from a previously reported study.^19^ The participants of that study, recruited from the San Francisco Bay Area, had no previous diagnosis of diabetes.^19^ Among the study subjects (32 females and 25 males), 5, 14, and 38 individuals met their criteria of “type 2 diabetes” (HbA1c ≥ 6.5%, FBG ≥ 126 mg/dL, or 2-h glucose during 75-g OGTT ≥ 200 mg/d), “pre-diabetes” (HbA1c > 5.7% and < 6.5%, FBG 100–125 mg/dL, or 2-h glucose during 75-g OGTT 140–199 mg/dL) and “normoglycemia” (glucose-related parameters below the diagnostic thresholds for pre-diabetes).^19^

### CGM-derived parameters

CGM_Mean and CGM_Std represent the mean and standard deviation of glucose values measured by CGM, respectively. CONGA, LI, JINDEX, HBGI, GRADE, MODD, MAGE, ADRR, MVALUE, and MAG were calculated using EasyGV software.^25^ Here, we introduced novel indices, the mean (AC_Mean) and the variance (AC_Var) of the autocorrelation function of glucose values at lags 1–30 with a lag of 5 min. While autocorrelation function has been used to analyze this type of time series data,^26,27^ it has not been thoroughly investigated to date. AC_Mean and AC_Var used in analyzing blood glucose levels after consuming standardized meals^19^ were calculated from the autocorrelation function at lags 1–10, as we had CGM data available for only 2.5 h after standardized meals were consumed.

DTW_Low, DTW_Mod, and DTW_Sev are previously proposed CGM-derived indices that represent the dysregulation of glycemia.^19^ These indices were calculated by a previously reported method.^19^ In brief, the time series data of CGM were fragmented into sliding windows of 2.5 h, with a 75% overlap. Then, by applying spectral clustering, three clusters of glucose patterns (low, moderate, and severe) were identified, and the fraction of time in each category was defined as DTW_Low, DTW_Mod, and DTW_Sev, respectively.

### Calculation of clinical indices

Insulinogenic index (I.I):

Ratio of the increment of immunoreactive insulin (IRI) to that of plasma glucose at 30 min after the onset of the OGTT.

Composite index:

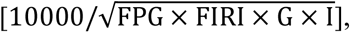

where FPG, FIRI, G, and I are fasting plasma glucose, fasting IRI, mean blood glucose levels, and mean serum IRI concentrations during the OGTT, respectively.

Oral DI:

Product of the composite index and the ratio of the area under the insulin concentration curve from 0 to 120 min to that for plasma glucose from 0 to 120 min, without using the data measured at 90 min, in the OGTT.

AUC_IRI:

Incremental area under the IRI concentration curve from 0 to 10 min during the hyperglycemic clamp.

Insulin sensitivity index (ISI):

The mean glucose infusion rate during the final 30 min of the clamp (mg/kg/min) divided by both the plasma glucose (mg/dL) and serum insulin (μU/mL) levels at the end of the clamp and then multiplying the resulting value by 100.

Clamp DI:

The product of AUC_IRI and ISI.

Metabolic clearance rate of inulin (MCRI):

Ratio of insulin infusion rate to the steady-state plasma insulin concentration during the hyperinsulinemic-euglycemic clamp test.

### Mathematical model estimating insulin sensitivity, secretion, and clearance

To estimate insulin sensitivity, insulin secretion, and insulin clearance from clamp tests, we constructed a mathematical model of the feedback loop that links glucose and insulin as shown in a previous study^28^ as follows:

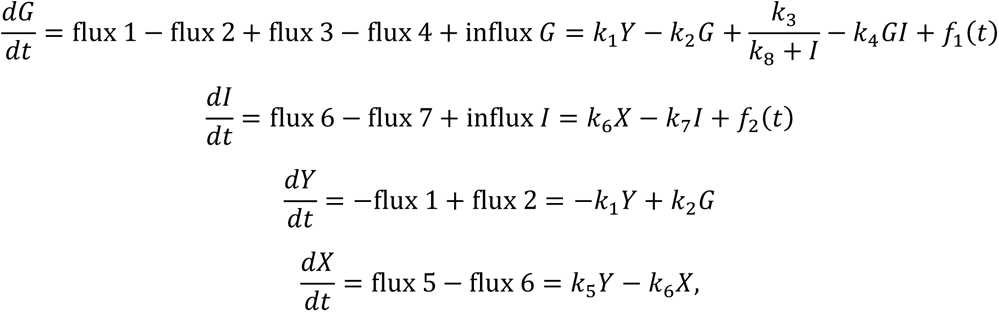

where the variables *G* and *I* denote blood glucose and insulin concentrations, respectively. The fluxes, influx *G* and influx *I* denote glucose and insulin infusions, respectively. These fluxes were estimated using a previously reported method.

For each of the 64 subjects, the parameters of the model to reproduce the time course were estimated by a meta-evolutionary programming method to search the minimum globally, followed by application of the nonlinear least squares technique to search the minimum locally, as previously described.^29^ Each parameter of the model for serum glucose and insulin concentration was estimated in the range from 10^−4^ to 10^4^. For these methods, the parameters were estimated to minimize the objective function value, which is defined as residual sum of the square (RSS) between the actual time course obtained by clamp analyses and the model trajectories. RSS used in the model for serum glucose and insulin concentration was given by the following equation:

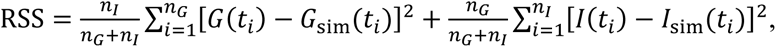

where *n*_*G*_ and *n*_*I*_ are the total numbers of time points of measuring blood glucose and insulin, respectively, and *t*_*i*_ is the time of i-th time point. *G*(*t*) is the time-averaged blood glucose concentration within the time range (*t* − 5) min to *t* min with every 1-min interval, *I*(*t*) is the blood insulin concentration at *t* min. *G*_sim_(*t*) and *I*_sim_(*t*) are simulated blood glucose and insulin concentrations, respectively. Blood glucose and insulin concentrations of each subject were normalized by dividing them by the respective maximum value. The numbers of parents and generations in the meta-evolutionary programming were 400 and 4000, respectively.

### Mathematical model used for simulating the characteristics of AC_Mean and AC_Var

In simulating the characteristics of AC_Mean and AC_Var, we used a simple and stable model,^30^ which can be written as follows:

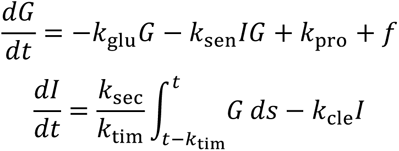

where the variables *G* and *I* denote blood glucose and insulin concentrations, respectively. Parameter values reported as the averages for healthy subjects were as follows:^30^

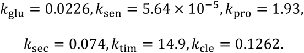

We simulated how 24-h profiles of *G* changed as *k*_sec_*k*_sen_ and *k*_cle_, which correspond to the DI and insulin clearance, respectively, were changed from one-half to twice as large as the values. Five mg/dL/min glucose was applied for 10 min at 6-, 12-, and 18-h as the external input of glucose *f*. We also calculated the AC_Mean and AC_Var from *G* added with zero-mean gaussian white noise with variances of 0.25, 0.5, or 1.

### Prediction models and statistical analyses

In this study, we investigated the predictive performance of CGM-derived indices for assessing the decline in blood glucose control ability across five main methodologies: multiple linear regression, partial least squares (PLS) regression, least absolute shrinkage operator (LASSO) regression, random forests, and logistic regression. Of note, these prediction models were conducted as post hoc analyses. The input variables for these models consisted of 27 variables: body mass index (BMI), abdomen circumference (ACir), body fat percentage, systolic blood pressure (SBP), diastolic blood pressure (DBP), total cholesterol (TC), triglycerides (TGs), low-density lipoprotein cholesterol (LDL-C), high-density lipoprotein cholesterol (HDL-C), FBG, HbA1c, CGM_Mean, CGM_Std, CONGA, LI, JINDEX, HBGI, GRADE, MODD, MAGE, ADRR, MVALUE, MAG, DTW_Mod, DTW_Sev, AC_Mean, and AC_Var. This modeling was conducted using scikit-learn, a python-based tool kit.

The predictive performance assessment of multiple linear regression included measures such as the coefficient of determination (*R*^2^), the adjusted coefficient of determination (Adj *R*^2^), and Akaike information criterion (AIC). The multicollinearity of the input variables was estimated by the variance inflation factor (VIF). PLS regression was conducted to estimate the importance of the input variables in predicting the DI. The variable importance in projection (VIP) scores,^31^ which were generated from PLS regression, were used for estimating the importance of the input variables. Lasso regression is a kind of linear regression with L1 regularization.^32,33^ The optimal regularization coefficients, lambda, were based on leave-one-out cross validation. For multiple linear regression, PLS regression, LASSO regression, and logistic regression, z-score normalization was performed on each input variable.

Random forest is an ensemble learning method, which generates classification decision trees by selecting subsets of input predictor variables randomly.^34^ The study employed 300 decision trees with Gini as the criterion for determining the best splits. The predictive performance of random forests was assessed using accuracy and F1 score based on leave-one-out cross-validation. The importance of the input variables in predicting glycemic anomaly is based on the permutation and the feature importance function of the random forest function. Boruta^35^ was also used to test whether the input variables is usable for the prediction.

Associations between indices were assessed using Spearman’s correlation test, and correlation coefficients were reported with 95% confidence intervals (Cis) through bootstrap resampling. The number of resamples performed to form the bootstrap distribution was set at 10000. *P* < 0.05 was considered statistically significant. Benjamini-Hochberg’s multiple comparison test was also performed with a significance threshold of Q < 0.05.

Hierarchical clustering analysis was also conducted using a method that combines Euclidean distance measure and Ward linkage. It was adopted after Z score normalization. Comparisons among individuals in each cluster were performed by analysis of variance followed by Tukey’s honestly significant difference test.

## RESULTS

### Autocorrelation of glucose levels fluctuations

In this study, our primary objective was to devise novel CGM-derived metrics that represent the glucose handling capacity of the living body. To achieve this, we conducted CGM, an OGTT, and a consecutive hyperglycemic and hyperinsulinemic-euglycemic clamp test in individuals without prior diabetes diagnosis, enabling us to focus on early indicators of reduced blood glucose control abilities.

We focused on the autocorrelation of glycemic fluctuation pattern, in particular, the mean (AC_Mean) and the variance (AC_Var) of the autocorrelation function at lags 1–30 with the length of the lag for 5 min, given that glucose levels were measured every 5 min with the CGM device (Figs. 1 and S2). Figure 1 shows the autocorrelation function of glycemic fluctuation pattern in two representative subjects (Subjects #13 and #46), both of whom were categorized as normal glucose tolerance (NGT), but had different clamp DI values: 65.7 for Subject #13 and 11.5 for Subject #46.

**Fig. 1.**
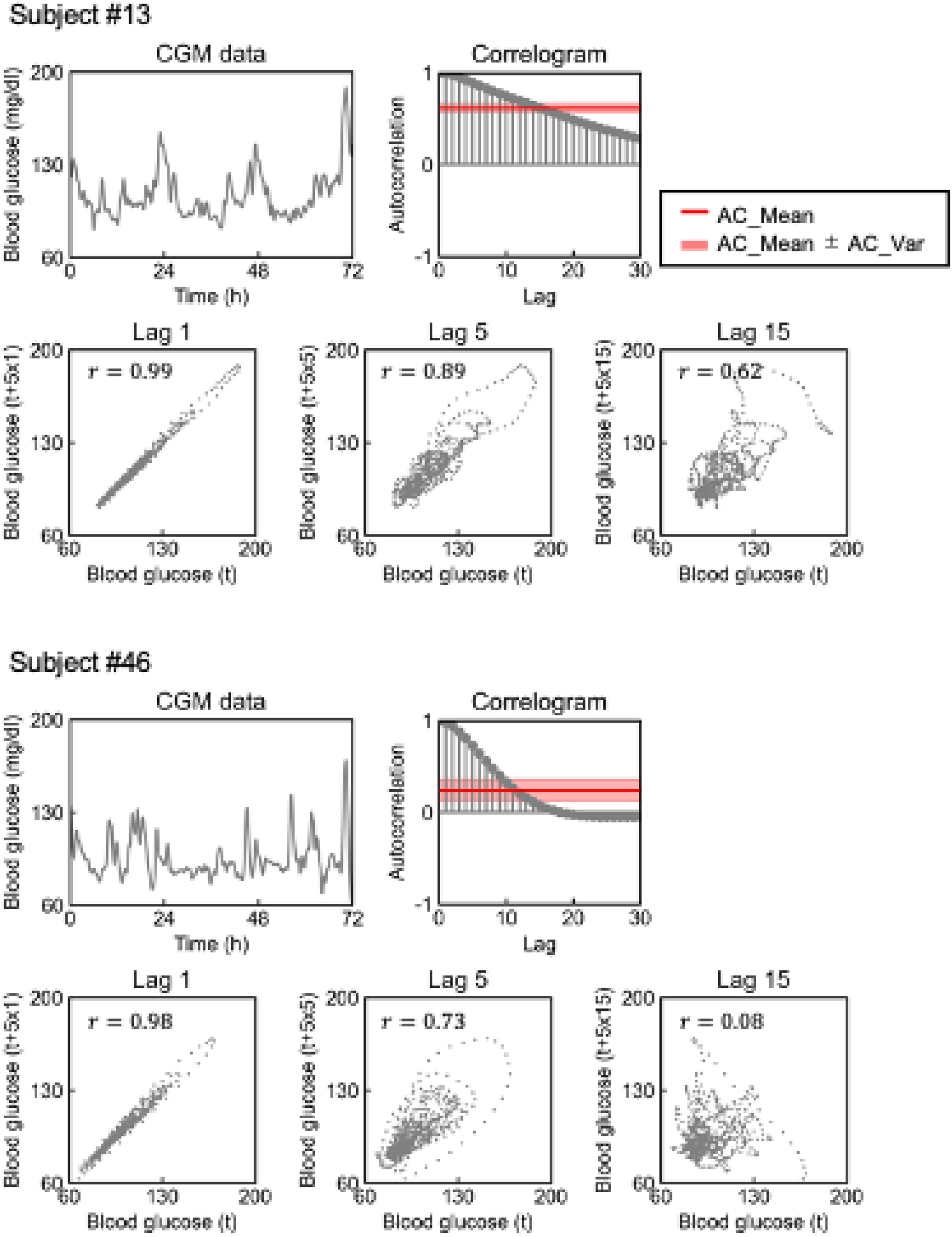
Blood glucose fluctuations and correlogram of the time series data. Time courses of blood glucose levels measured by continuous glucose monitoring in two subjects and their correlograms. Red lines indicate the mean value of the autocorrelation function (AC_Mean), and red shaded areas indicate the variance value of the autocorrelation function (AC_Var) (see Methods). Lag 1 autocorrelation of blood glucose measurements taken at 5-min intervals refers to the relationship between the current value (Blood glucose [t]) and the value recorded one time point later (Blood glucose [t+5x1]). Lag 5 and lag 15 similarly refers to the relationship between the current value (blood glucose [t]) and the value recorded five time points later (blood glucose [t+5x5]) and 15 time points later (blood glucose [t+5x15]), respectively.

For individuals with slower fluctuations in blood glucose levels, such as Subject #13, the autocorrelation function displayed a slower rate of decrease with increasing lag (AC_Mean: 0.62, AC_Var: 0.049). By contrast, subjects with more rapid glucose fluctuations, such as Subject #46, exhibited a relatively swift decrease in autocorrelation function as the lag increased (AC_Mean: 0.24, AC_Var: 0.12). It is noteworthy that although Subject #46 had a lower clamp DI than Subject #13, the mean (CGM_Mean) and standard deviation (CGM_Std) of glucose values were also lower in Subject #46 (CGM_Mean: 95; CGM_Std: 16) than in Subject #13 (CGM_Mean: 107; CGM_Std: 18). Moreover, given the definition formula for the autocorrelation function, the autocorrelation function would not change if calculated from blood glucose levels standardized to a mean of 0 and a variance of 1, indicating a possibility that AC_Mean and AC_Var can possess characteristics that differ from the average or the variability of glucose levels.

Given these findings, we hypothesized that these indices might possess some information about underlying glucose regulatory abilities that are not discernible through conventional measures such as OGTT or the mean and standard deviation of blood glucose levels. Consequently, our primary research objective was to ascertain the association between AC_Mean and AC_Var and glucose handling capacities, such as the DI and insulin clearance. Of note, the DI and insulin clearance reportedly reflect glucose dysregulation well and predict the development of future T2DM beyond FBG and the plasma glucose concentration at 120 min during the OGTT (PG120).^36,37^

### Relationship between indices from CGM and those from OGTT or clamp tests

We compared AC_Mean and AC_Var with indices obtained from OGTT or clamp tests to thoroughly assess the potential of these indices in estimating a decline in blood glucose control ability (Figs. 2A and S3). In this study, CGM, OGTT, and clamp tests were performed on 52 NGT, 9 impaired glucose tolerance (IGT), and 3 T2DM individuals (Table S1). PG120, I.I., composite index, and oral DI were calculated from the OGTT. AUC_IRI, ISI, and clamp DI were calculated from clamp tests. Additionally, *k*_4_, *k*_5_, and *k*_7_ were calculated using a previously established mathematical model.^29^ PG120 is the plasma glucose concentration at 120 min during OGTT. I.I., AUC_IRI, and *k*_5_ correspond to insulin secretion. Composite index, ISI, and *k*_4_ correspond to insulin sensitivity. *k*_7_corresponds to insulin clearance. DI is the product of insulin sensitivity and insulin secretion. Of note, we refrained from employing multiple testing corrections encompassing all pairwise comparisons in Figure 2A due to the interdependent nature of the conducted tests; we analyzed various indicators that represent the same abilities, aiming to confirm the meaningful correlations observed.

**Fig. 2.**
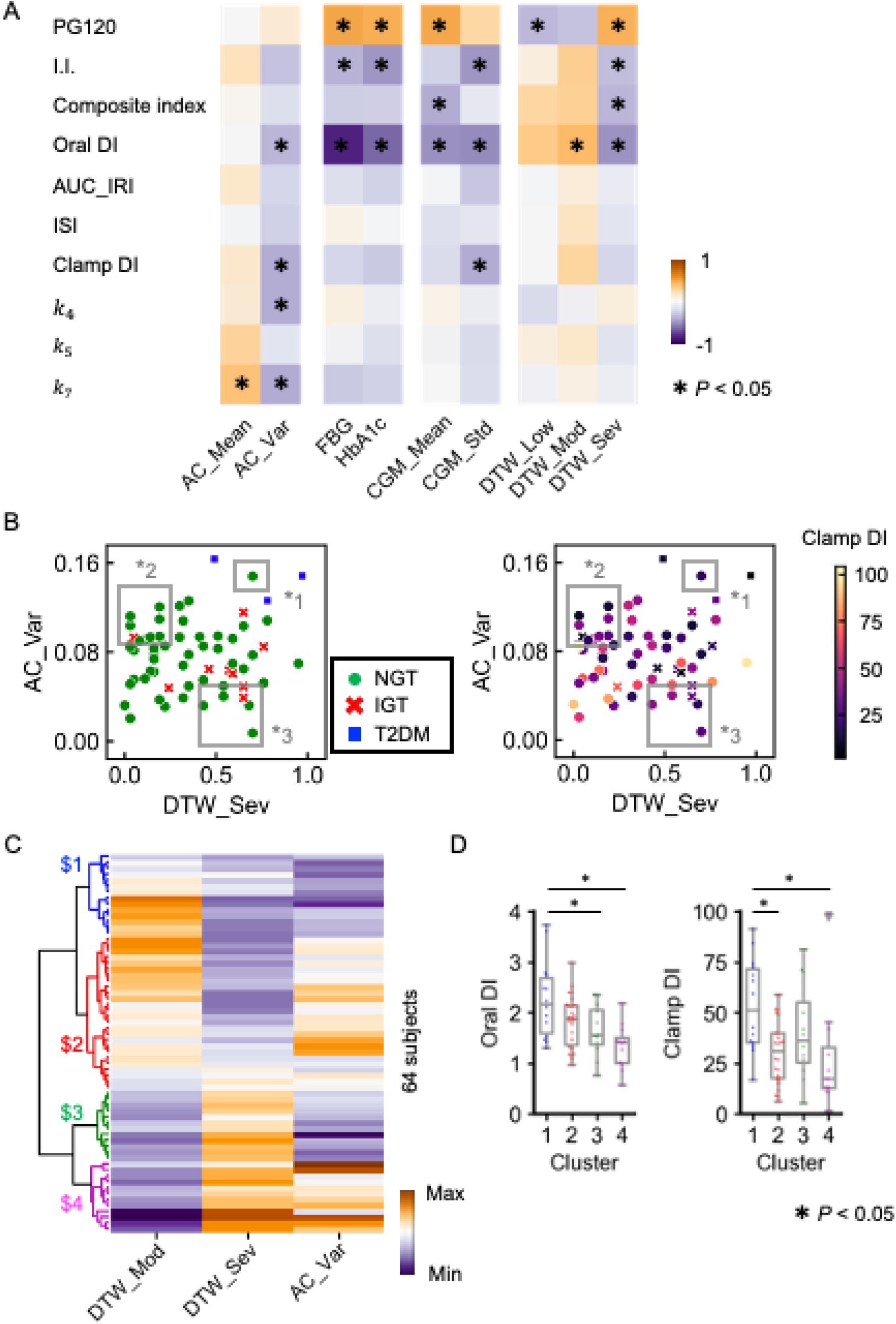
Correlation between the indices from CGM and those from OGTT or clamp tests. (A) Heatmap of Spearman’s correlation coefficient, with the *P* values corresponding to testing the hypothesis of no correlation. CGM_Mean and CGM_Std indicate the mean value and standard deviation of blood glucose levels measured by CGM, respectively. DTW_Low, DTW_Mod, and DTW_Sev are previously reported indices of glucose variability and glucose dysregulation^19^ (see Methods). AC_Mean and AC_Var are the mean and the variance value of the autocorrelation function of blood glucose fluctuations measured by CGM, respectively. The parameters *k*_4_, *k*_5_, and *k*_7_ correspond to insulin sensitivity, insulin secretion, and insulin clearance, respectively. (B) Scatter plots for DTW_Sev versus AC_Var. Subjects were colored based on diabetes diagnosis (the left) and the value of clamp DI (the right). This diagnosis is based on the American Diabetes Association Guidelines of HbA1c, fasting blood glucose, and OGTT. An NGT subject who had high AC_Var and high DTW_Sev had low clamp DI (*1). Some subjects had low DTW_Sev and low clamp DI (*2), and others had low AC_Var and low clamp DI (*3). (C) Hierarchical clustering analysis of the standardized CGM-derived indices using Euclidean distance as a metric with the Ward method. The columns represent the standardized value of DTW_Mod, DTW_Sev, and AC_Var. The rows represent individual subjects. The subjects are grouped and sorted according to their degree of relatedness. (D) Box plots of the value of oral DI and clamp DI for each cluster. Blue, red, green, and magenta are related to clusters 1, 2, 3, and 4, respectively. The boxes denote the median and upper and lower quartiles. Each point corresponds to the value for a single subject. \**P* < 0.05. Abbreviations: AUC_IRI, area under insulin curve during the first 10 min of hyperglycemic clamp test; CGM, continuous glucose monitoring; DI, disposition index; I.I., insulinogenic index; ISI, insulin sensitivity index; NGT, normal glucose tolerance; OGTT, oral glucose tolerance test; PG120, plasma glucose concentration at 120 min during the OGTT.

AC_Mean exhibited a statistically significant correlation with *k*_7_ (r = 0.28; 95% CI: 0.04 to 0.50). AC_Var displayed statistically significant correlations with the oral DI (r = – 0.28; 95% CI: –0.51 to –0.02), clamp DI (r = –0.31; 95% CI: –0.52 to –0.07), *k*_4_ (r = –0.31; 95% CI: –0.52 to –0.06), and *k*_7_ (r = –0.31; 95% CI: –0.54 to –0.06). Since we were testing two indices, AC_Mean and AC_Var, we also applied the Benjamini-Hochberg’s multiple comparison test for each comparison, resulting in Q < 0.05 for these comparisons, validating the significance of the identified relationships among these indices. To further validate the relationships among these indices, we also investigated the relationships among AC_Mean, AC_Var, *k*_4_*k*_5_ (corresponding to the clamp DI) and MCRI (corresponding to insulin clearance). AC_Mean exhibited a statistically significant correlation with *k*_4_*k*_5_ (r = 0.29; 95% CI: 0.05 to 0.50). AC_Var displayed statistically significant correlations with *k*_4_*k*_5_ (r = –0.29; 95% CI: –0.50 to –0.05) and MCRI (r = –0.33; 95% CI: –0.55 to –0.08). Additionally, AC_Mean and AC_Var calculated from different lags were also significantly correlated with *k*_7_, oral DI, and clamp DI (Fig. S4). Collectively, we conclude that AC_Mean and AC_Var are associated with the DI or insulin clearance.

For comparison, we also assessed commonly used indicators such as FBG and HbA1c, which have been recommended for pre-DM screening.^9–11^ Moreover, we examined CGM-derived basic features such as mean (CGM_Mean) and standard deviation (CGM_Std) as well as CGM-derived indices, DTW_Low, DTW_Mod, and DTW_Sev. These indices have been shown to identify populations with decreased ability to control blood glucose within populations primarily composed of individuals categorized as having NGT.^19^ DTW_Low, DTW_Mod, and DTW_Sev were calculated using a previously reported method:^19^ low DTW_Low and high DTW_Sev indicate high glucose concentration and variability and glucose dysregulation.

The indices that exhibited significant correlations with AC_Mean or AC_Var differed from those significantly correlated with FBG, HbA1c, or other indices derived from CGM data; the other indices were not significantly correlated with all of the oral DI, clamp DI, *k*_4_, and *k*_7_. By contrast, FBG, HbA1c, or the other CGM-derived indices were significantly correlated with PG120, I.I., and the composite index, where AC_Mean and AC_Var exhibited no significant correlations.

### Estimation of the decreased ability to control blood glucose by combining CGM-derived indices

Based on these findings, we hypothesized that a more accurate identification of individuals with disrupted glucose regulation could be achieved by combining AC_Mean and AC_Var with conventional indices. To test this hypothesis, we investigated the relationship among AC_Var, DTW_Sev, diabetes diagnosis, and clamp DI – an indicator of glycemic disability and a predictor of T2DM^38^ (Fig. 2B). Due to a significant correlation between AC_Mean and AC_Var (r = –0.74; 95% CI: –0.85 to –0.60) (Fig. S3), we focused on AC_Var. AC_Var was not significantly correlated with DTW_Sev (r = 0.06; 95% CI: –0.20 to 0.32). In subjects with T2DM, both AC_Var and DTW_Sev were relatively high. Some subjects with high values of AC_Var and DTW_Sev were diagnosed with NGT or IGT, but their clamp DI was relatively low (Fig. 2B, *1). Some subjects had low DTW_Sev and low clamp DI (Fig. 2B, *2), and others had low AC_Var and low clamp DI (Fig. 2B, *3). In these subjects, either AC_Var or DTW_Sev was relatively high, suggesting a potential for combined use to enhance accuracy in identifying individuals with impaired glucose control.

Theoretically, dynamic time warping, an algorithm that is employed to calculate DTW_Sev, aligns time series data globally and may not fully account for autocorrelated structure information.^39^ Consequently, DTW_Sev is mainly affected by the mean and the variance of glucose levels.^40,41^ On the other hand, AC_Var is calculated from autocorrelation of glucose levels and autocorrelation is not affected by the mean and the variance of levels given its definition formula. Collectively, these rationales also indicate that DTW_Sev and AC_Var can have different aspects of glucose characteristics and that a combination of these two indices may be useful in estimating glucose handling capacities.

To further explore the efficacy of this combination, we conducted clustering analysis based on these indices (Fig. 2C). The sum of DTW_Low, DTW_Mod, and DTW_Sev was 1, DTW_Low was excluded from this analysis. We divided the subjects into four groups based on the relatedness of the indices: cluster $1 mainly contained subjects with low DTW_Sev and low AC_Var, cluster $2 mainly contained subjects with low DTW_Sev and relatively high AC_Var, cluster $3 mainly contained subjects with relatively high DTW_Sev and low AC_Var, and cluster $4 mainly contained subjects with relatively high DTW_Sev and relatively high AC_Var. Subjects in cluster $3 had statistically significantly lower oral DI than those in cluster $1 (Fig. 2D), and subjects in cluster $2 had statistically significantly lower clamp DI than those in cluster $1, suggesting that subjects with high DTW_Sev (cluster $3) or high AC_Var (cluster $2) have glycemic disabilities. As expected, subjects in cluster $4 had statistically significantly lower oral DI and clamp DI than those in cluster $1 (Fig. 2D).

We also conducted multiple regression analyses among oral DI, clamp DI, and the CGM-derived indices (Table 1). *R*^2^ of the models that predicted oral DI and clamp DI from DTW_Mod, DTW_Sev, and AC_Var were 0.24 and 0.14, respectively. AC_Var had an independent negative correlation with oral DI and clamp DI that was statistically significant (*P* = 0.029 and *P* = 0.010, respectively), suggesting that AC_Var contributes to the prediction of DI independently of DTW_Mod and DTW_Sev. AC_Var also had a negative correlation with clamp DI that was statistically significant (*P* = 0.048) independently of CGM_Std (Table 1C), which was also significantly correlated with clamp DI (Fig. 2A). *R*^2^, the adjusted coefficient of determination (Adj *R*^2^), and the AIC of the model that predicted clamp DI from CGM_Std and AC_Var were 0.18, 0.15, and 583, respectively. Of note, *R*^2^, Adj *R*^2^, and AIC of the model that predicted clamp DI from oral DI were only 0.15, 0.14, and 583, respectively. Collectively, we conclude that combining AC_Var with conventional CGM-derived indices can increase the accuracy of predicting a decreased ability to control blood glucose.

**Table 1.** Multiple regression analyses with oral DI index or clamp DI. (A) Multiple regression analysis between oral DI and CGM-derived indices, DTW_Mod, DTW_Sev, and AC_Var. (B) Multiple regression analysis between clamp DI and CGM-derived indices, DTW_Mod, DTW_Sev, and AC_Var. (C) Multiple regression analysis between clamp DI and CGM-derived indices, CGM_Std and AC_Var. \**P* < 0.05

|  | Coefficient | SE | *P* | 95% CI |
| --- | --- | --- | --- | --- |
| DTW_Mod | 0.029 | 0.17 | 0.87 | -0.31 - 0.37 |
| DTW_Sev | -0.22 | 0.17 | 0.20 | -0.56 - 0.12 |
| AC_Var | -0.16 | 0.071 | 0.029* | -0.3 - -0.017 |

(B) Multiple regression analysis between clamp DI and CGM-derived indices, DTW\_Mod, DTW\_Sev, and AC\_Var.
|  | Coefficient | SE | *P* | 95% CI |
| --- | --- | --- | --- | --- |
| DTW_Mod | 7.7 | 7.1 | 0.28 | -6.4 - 22 |
| DTW_Sev | 4.6 | 7.1 | 0.52 | -9.6 - 19 |
| AC_Var | -7.8 | 2.9 | 0.010* | -14 - -1.9 |

(C) Multiple regression analysis between clamp DI and CGM-derived indices, CGM\_Std and AC\_Var. \**P* < 0.05
|  | Coefficient | SE | *P* | 95% CI |
| --- | --- | --- | --- | --- |
| CGM_Std | -6.5 | 3.0 | 0.035* | -12.5 - -0.48 |
| AC_Var | -6.1 | 3.0 | 0.048* | -12 - -0.05 |

DI, disposition index; CGM, continuous glucose monitoring.

### Predicting DI from CGM, single-point blood tests, and physical measurement-derived indices

To predict DI, which reflects glycemic disability and has been suggested as a predictor of the development of T2DM,^37^ we conducted multiple linear regression analyses. Since the purpose of this study was to estimate glucose intolerance from relatively easy-to-measure indices, we included only CGM-derived indices, indices from a single blood test, and those from physical measurements as the input variables, as follows: CGM_Mean, CGM_Std, CONGA, LI, JINDEX, HBGI, GRADE, MODD, MAGE, ADRR, MVALUE, MAG, DTW_Mod, DTW_Sev, AC_Mean, AC_Var, TC, TGs, LDL-C, HDL-C, FBG, HbA1c, BMI, ACir, body fat percentage, SBP, and DBP.

To assess the multicollinearity of the input variables, we investigated the VIF of these variables (Fig. 3A). We removed the variable with the highest VIF one by one until the VIF of all variables were less than 10 (Fig. 3B), resulting in 18 variables (DBP, SBP, AC_Var, AC_Mean, HBGI, MAG, GRADE, DTW_Mod, ACir, BMI, MODD, FBG, HbA1c, TG, HDL-C, LDL-C, body fat percentage, MVALUE). AC_Mean and AC_Var were included in these 18 variables, suggesting their relatively low multicollinearity with other indices. *R*^2^ of the models that predicted oral DI and clamp DI from the 18 variables were 0.64 and 0.50, respectively (Fig. S5A, B). Adj *R*^2^ and AIC of the models that predicted oral DI (Fig. S5A) and clamp DI (Fig. S5B) using both CGM-derived indices and indices from a single-point blood test or physical measurements were better than those of models that used only CGM-derived indices (Figs. S6A, B) or a single-point blood test and physical measurement-derived indices (Fig. S6C, D), indicating that CGM-derived indices in addition to non-CGM indices contribute to a more accurate DI prediction.

**Fig. 3.**
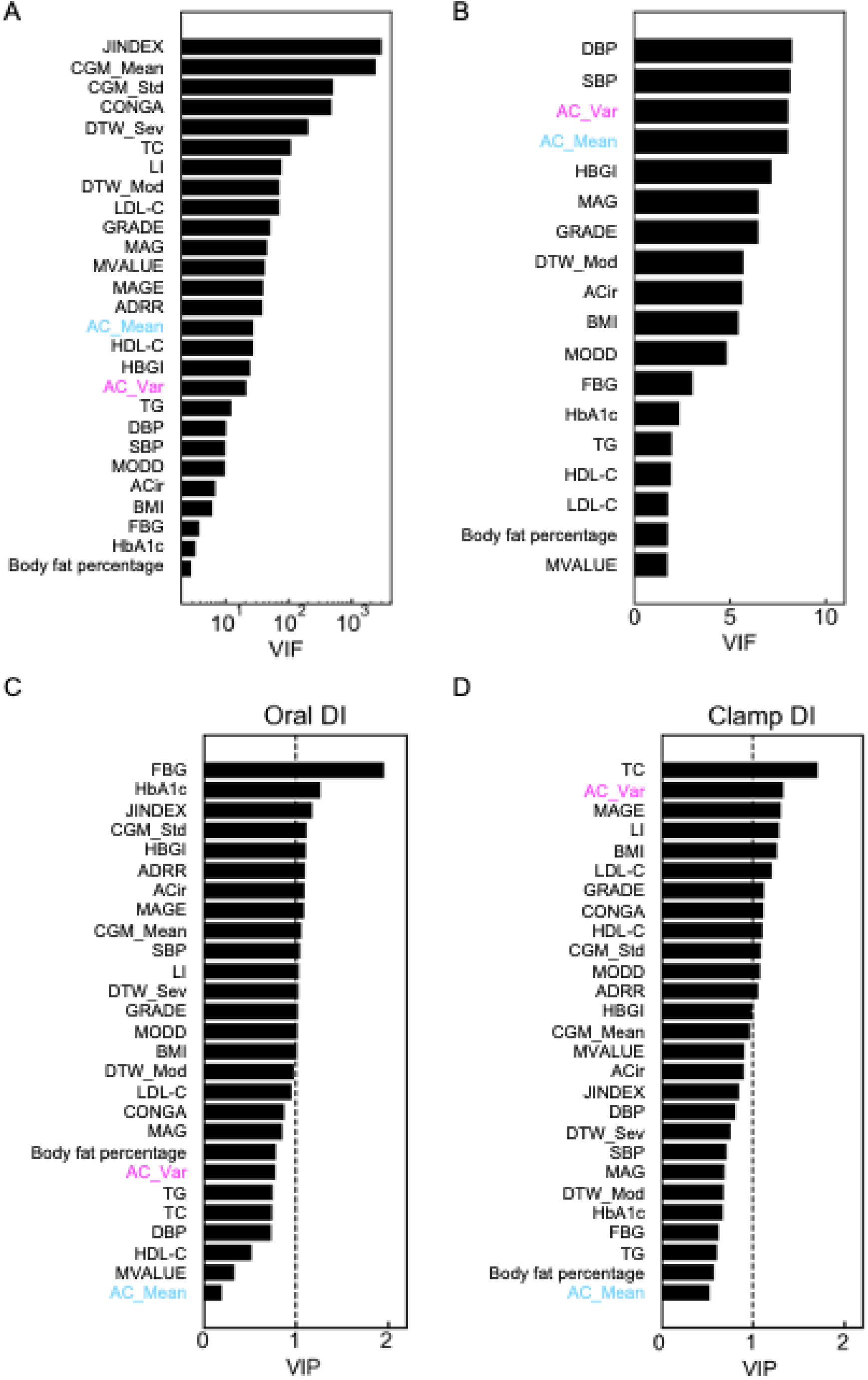
Multivariate analyses for predicting the DI. (A) VIF of all variables. (B) VIF of each variable remaining after removing the variable with the highest VIF one by one until the VIF of all variables are less than 10. (C) VIP generated from the PLS regression predicting oral DI. Variables with a VIP ≥ 1 (the dotted line) were considered to be significantly contributed to the prediction. (D) VIP generated from the PLS regression predicting clamp DI. Abbreviations: ACir, abdomen circumference; BMI, body mass index; DBP, diastolic blood pressure; DI, disposition index; FBG, fasting blood glucose; HDL-C, high-density lipoprotein cholesterol; LDL-C, low-density lipoprotein cholesterol; PLS: partial least squares; SBP, systolic blood pressure; TC, total cholesterol; TG, triglycerides; VIF, variance inflation factor; VIP, variable importance in projection.

To avoid overfitting and to investigate the importance of the input variables in predicting DI, we also applied machine learning models, including PLS regression with VIP scores^31^ (Fig. 3C, D) and least absolute shrinkage and selection operator (Lasso) regression^33^ (Fig. S7). These regression models have been used for studies where the number of input variables are large relative to the sample size.^42,43^ PLS regression has been used for datasets with mutually correlated input variables and output variable, and important predictors can be estimated by VIP.^31^ Here, high VIP indicates high contribution of the variables in predicting DI. Lasso employs L1 regularization, which leads to models with fewer parameters, and has been used to select useful features to predict DM from numerous input variables.^32^

The cross validation indicated that the optimal number of PLS components was 2. At the component, the VIP of some CGM-derived indices, including AC_Var, was higher than 1 (Fig. 3D). The leave-one-out cross validation indicated that the optimal regularization coefficients of lasso (Lambda) were 0.061 for oral DI and 3.49 for clamp DI (Fig. S7A, B). At these Lambdas, the coefficients of AC_Var were estimated as non-zero coefficients for both oral DI and clamp DI (Fig. S7C–F). Collectively, these results indicate that CGM-derived indices, including AC_Var, contribute to the prediction of DI.

We also investigated the characteristics of AC_Mean and AC_Var in predicting the decreased ability to control blood glucose by conducting random forests and a logistic regression analysis with L1 regularization (Supplementary text 2, Figs. S8–10), suggesting that including AC_Mean or AC_Var alongside conventional indices enhances the accuracy of predicting decreased glucose control abilities.

### Relationship among clinical parameters

To provide an overview of the relationship among indices derived from OGTT, clamp tests, CGM, and other clinical parameters from a single blood test or physical measurements, we constructed a correlation network (Fig. 4). Correlation networks have been used to elucidate the interdependency and interconnectivity among different biomarkers of complex metabolic disorders such as T2DM.^44^ In this network, AC_Mean and AC_Var were correlated with some insulin-related indices (blue nodes), but the correlations with other indices (red, magenta, and green nodes) were relatively small. We also applied the Benjamini-Hochberg’s multiple comparison test for each comparison, and showed relationships with Q < 0.05 in Table S2. AC_Var was significantly correlated with *k*_7_, oral DI, and clamp DI (Table S2). Collectively, we conclude that the autocorrelation function of glucose levels has the ability to capture a diminished capacity for blood glucose control that remains undetected by conventional indices.

**Fig. 4.**
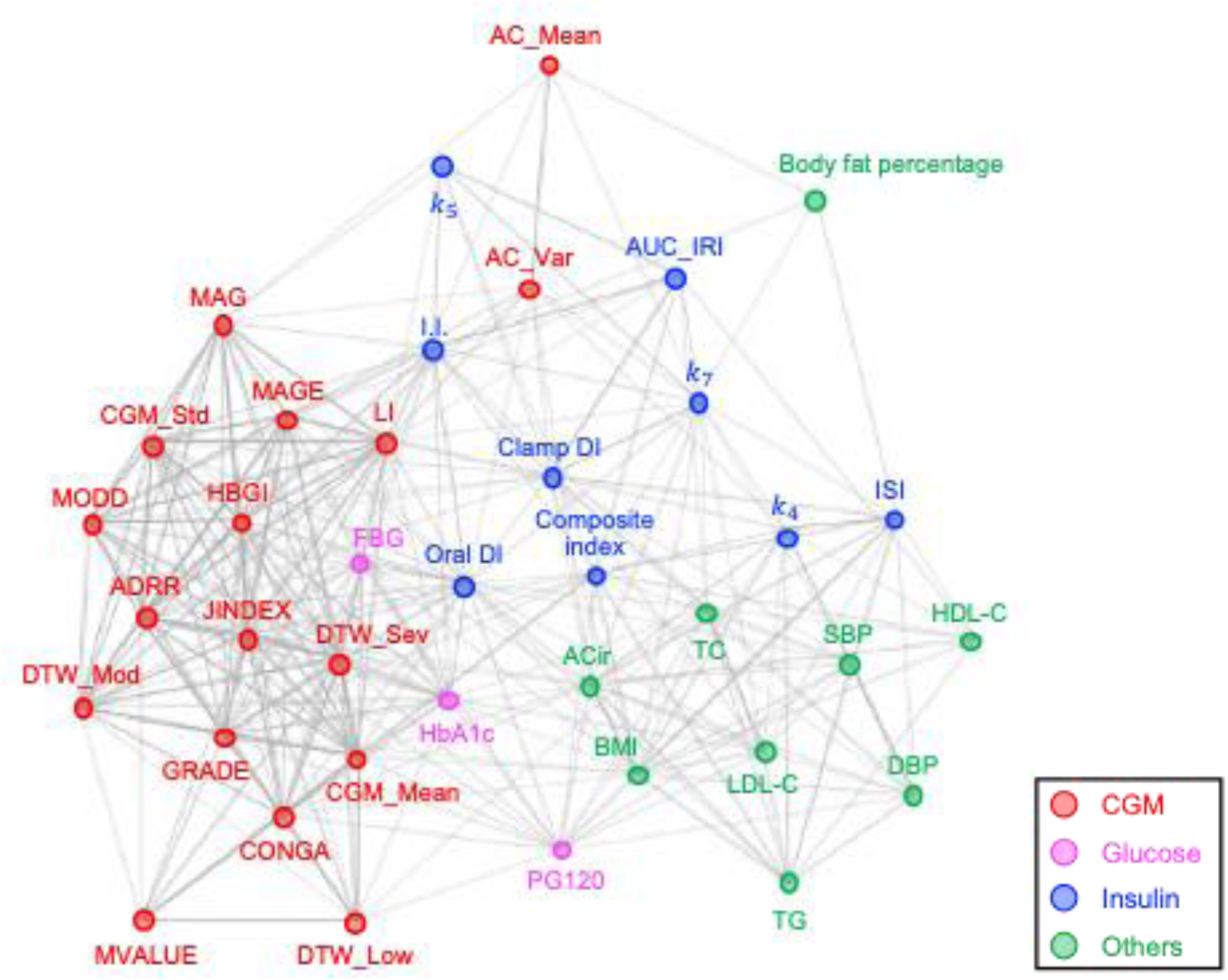
Correlation network of clinical parameters. A spring layout of the correlation network of 17 CGM-derived indices; 3 blood glucose level-related indices; 9 insulin sensitivity, secretion, and clearance-related indices; and 9 other lifestyle diseases-related indices obtained from a single blood test or physical measurement. Relationship with the absolute values of the Spearman’s correlation coefficient of 0.25 or higher are connected with edges. The width of the edges is proportional to the corresponding correlation coefficient. The color of the nodes indicates the type of indices: red, magenta, blue, green is associated with CGM, blood glucose levels, insulin, and other lifestyle-related diseases, respectively. Abbreviations: ACir, abdomen circumference; BMI, body mass index; CGM: continuous glucose monitoring; DBP, diastolic blood pressure; DI, disposition index; FBG, fasting blood glucose; HDL-C, high-density lipoprotein cholesterol; I.I., insulinogenic index; ISI, insulin sensitivity index; LDL-C, low-density lipoprotein cholesterol; oral AUC_IRI, area under insulin curve during the first 10 min of hyperglycemic clamp test; PG120, plasma glucose concentration at 120 min during the oral glucose tolerance test; SBP, systolic blood pressure; TC, total cholesterol; TGs, triglycerides. The parameters *k*_4_, *k*_5_, and *k*_7_ correspond to insulin sensitivity, insulin secretion, and insulin clearance, respectively.

### Validation of the contributions of AC_Mean and AC_Var in predicting a decreased ability to control blood glucose using an independent dataset

To further validate the significance of AC_Mean and AC_Var in predicting reduced blood glucose control capacity, we conducted multivariate analyses (Fig. 5A–D) and constructed a correlation network (Fig. 5E) by using a different dataset.^19^ This dataset included 57 participants who were free from prior diabetes diagnosis, with 5 individuals meeting the criteria for T2DM, 14 having pre-DM, and the remaining participants having NGT.

**Fig. 5.**
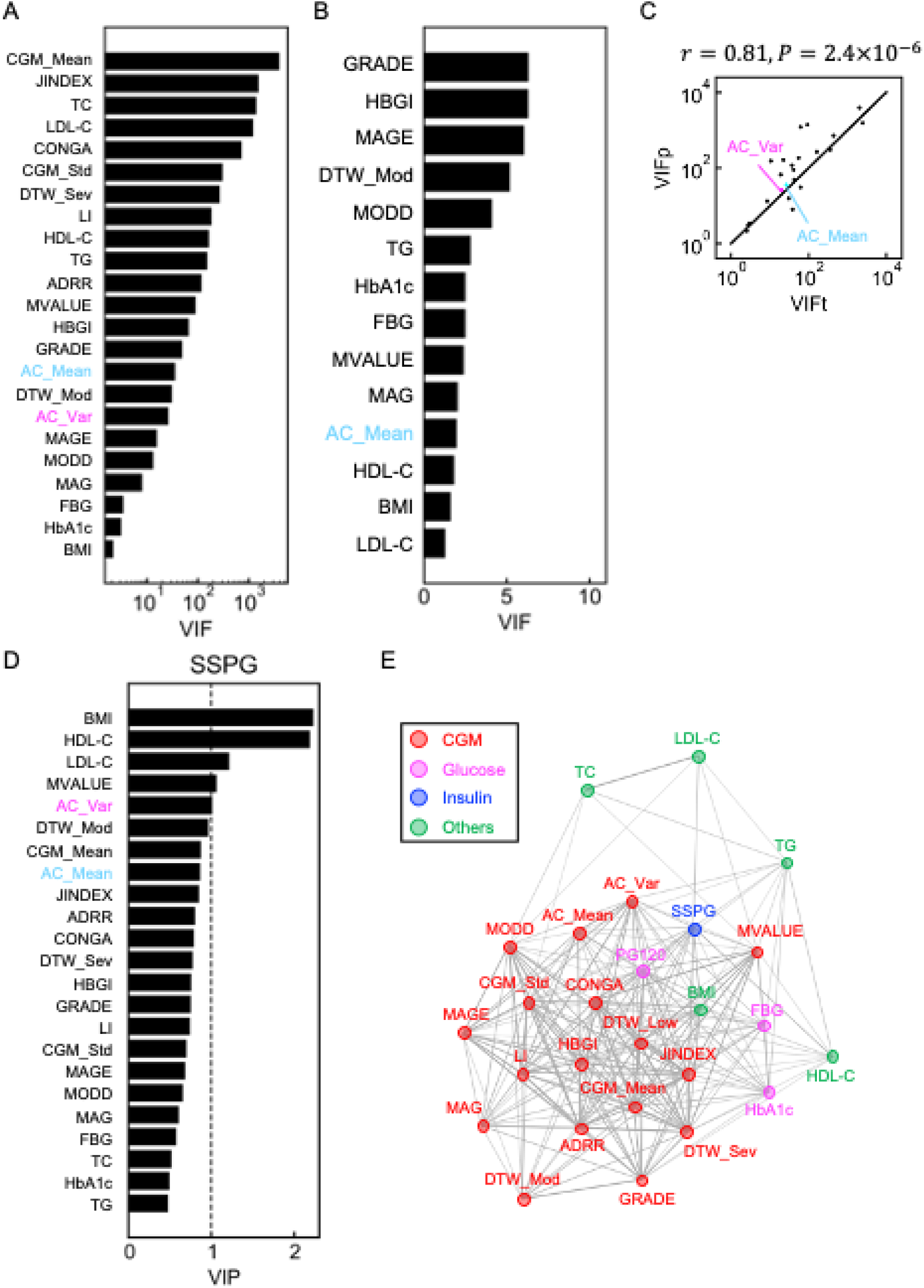
Validation of the characteristics of AC_Mean and AC_Var using a previously reported dataset. A previously reported dataset^19^ was used in this analysis. (A) VIF of all variables. (B) VIF of each variable remaining after removing the variable with the highest VIF one by one until the VIF of all variables are less than 10. (C) Scatter plot for the VIF of the indices calculated using the previously reported dataset (VIFp) versus those calculated using the dataset obtained in this study (VIFt). Each point corresponds to the values for an index. *r* is Spearman’s correlation coefficient and the *P* value is for testing the hypothesis of no correlation. (D) VIP generated from the PLS regression predicting SSPG. Variables with a VIP ≥ 1 (the dotted line) were considered to be significantly contributed to the prediction. (E) A spring layout the of correlation network of 17 CGM-derived indices, 3 blood glucose levels-related indices, 1 insulin sensitivity-related index, 5 other lifestyle diseases-related indices obtained from a single blood test or physical measurement. Relationships with the absolute values of the Spearman’s correlation coefficient of 0.25 or higher are connected with edges. The width of the edges is proportional to the corresponding correlation coefficient. The color of the nodes indicates the type of indices: red, magenta, blue, green is associated with CGM, blood glucose levels, insulin, and other lifestyle-related diseases, respectively. Abbreviations: BMI, body mass index; CGM, continuous glucose monitoring; FBG, fasting blood glucose; HDL-C, high-density lipoprotein cholesterol; LDL-C, low-density lipoprotein cholesterol; PLS: partial least squares; SSPG, steady state plasma glucose; TC, total cholesterol; TGs, triglycerides; VIF, variance inflation factor; VIP, variable importance in projection.

The assessment of multicollinearity using VIF revealed that AC_Mean exhibited relatively low multicollinearity with other indices (Fig. 5A, B), consistent with findings from previous analyses (Fig. 3A, B). The VIF of the indices calculated using the previously reported dataset (VIFp) and those calculated using the dataset obtained in this study (VIFt) were statistically significantly correlated (Fig. 5C), indicating the validity of the relationship among the clinical parameters. The cross validation indicated that the optimal number of components of the PLS regression model predicting SSPG, which indicates insulin sensitivity, was 2. At the component, VIP of AC_Var in predicting SSPG was higher than 1 (Fig. 5D), indicating that AC_Var contribute to the prediction of SSPG. To avoid overfitting and to investigate the contributions of the input variables in estimating SSPG, we also conducted Lasso (Fig. S11). The leave-one-out cross validation showed that the optimal regularization coefficient, Lambda, was 0.69 (Fig. S11A, B). At the Lambda, the coefficients of AC_Mean and AC_Var were estimated as non-zero coefficients (Fig. S11C), suggesting that AC_Mean and AC_Var contribute to the prediction of SSPG.

A correlation network showed that AC_Mean and AC_Var were significantly correlated with SSPG (r = –0.36; 95% CI: –0.57 to –0.09 and r = 0.48; 95% CI: 0.23 to 0.68, respectively), but the correlations with other CGM-derived indices were relatively modest (Fig. 5E). We also applied the Benjamini-Hochberg’s multiple comparison test for each comparison, and showed relationships with Q < 0.05 in Table S3. Both AC_Mean and AC_Var were significantly correlated with SSPG (Table S3). These results were consistent with the result that AC_Var was significantly correlated with *k*_4_, which corresponds to insulin sensitivity (Fig. 2A). These results confirm that the characteristics of AC_Mean and AC_Var are reproducible in capturing reduced blood glucose control ability.

To investigate the effects of meals on AC_Mean and AC_Var, we calculated the indices using the previously reported CGM data that were collected after consuming standardized meals^19^ (Fig. S12). The standardized meals were about the same in calories, but differed in the amounts of proteins, fat, and fiber: cornflakes and milk (Cereal) were low in fiber and high in sugar, peanut butter sandwiches (Bread and PB) were high in fat and high in protein, and PROBAR protein bars (Bar) were moderate in fat and protein, as previously described.^19^ The previously reported indices, DTW_Low, DTW_Mod, and DTW_Sev, were able to capture the differences in glucose fluctuations due to different meals (Fig. S12A).^19^ By contrast, one-way analysis of variance for testing the significance of differences in AC_Mean and AC_Var for each meal showed no significant difference (Fig. S12B), suggesting that AC_Mean and AC_Var were more robust to meal types.

### AC_Mean and AC_Var capture changes in blood glucose dynamics in the early stage of glucose intolerance

To further validate the predictive characteristics of AC_Mean and AC_Var in identifying reduced blood glucose control ability, we characterized them using simulated blood glucose data. We previously reported that as DI decreases, insulin clearance decreases simultaneously in the early stage of glucose intolerance.^28^ Hence, we evaluated how AC_Mean and AC_Var change as DI and insulin clearance decrease simultaneously.

For simulation of the mathematical model (see Methods), we used parameters reported as the mean values in NGT.^30^ We changed DI (*k*_sec_*k*_sen_) and insulin clearance (*k*_cle_) from one-half to twice the NGT’s values (Fig. 6A). As *k*_sec_*k*_sen_ and *k*_cle_ increased, FBG levels remained unchanged (Fig. 6A, #*α*), but there was a point at which blood glucose levels decreased (Fig. 6A, #*β*) and a point at which blood glucose levels increased (Fig. 6A, #*γ*). As the pattern of blood glucose dynamics changed in this way, AC_Mean increased and AC_Var decreased (Fig. 6B), consistent with the results that AC_Mean was positively correlated with insulin clearance, and AC_Var was negatively correlated with DI and insulin clearance (Fig. 2A).

**Fig. 6.**
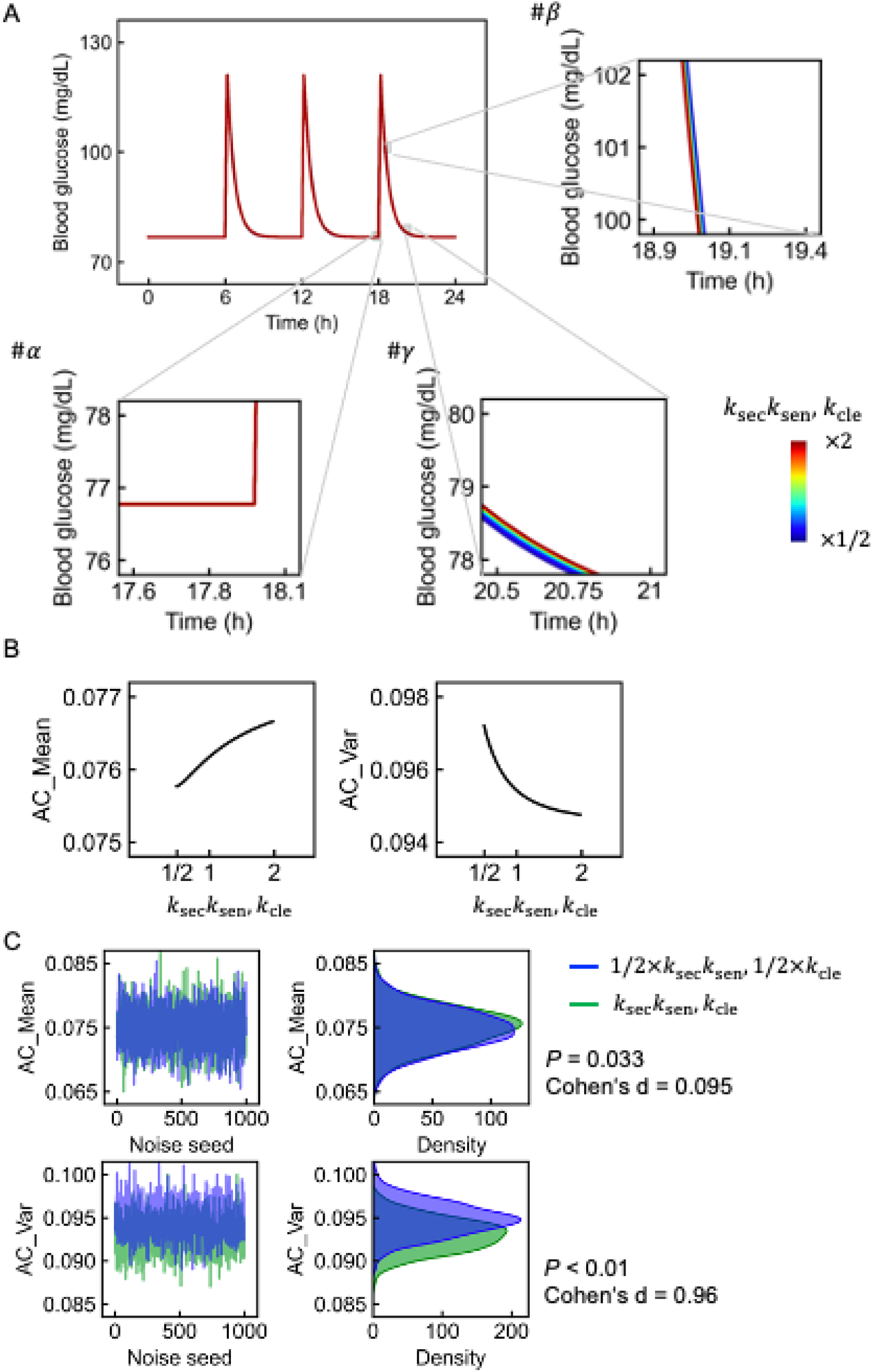
Characterization of AC_Mean and AC_Var using simulated blood glucose. (A) 24-h simulated glucose concentration. The color of the line is based on the values of *k*_sec_*k*_sen_ and *k*_cle_. The enlarged views of the areas are shown in #*α*-#*γ*. (B) The relationship between *k*_sec_*k*_sen_, *k*_cle_ and AC_Mean (the left). That of AC_Var is shown in the right. The horizontal axis represents the ratio of *k*_sec_*k*_sen_ and *k*_cle_ to the reported average values for healthy subjects. (C) AC_Mean (the upper panels) and AC_Var (the lower panels) simulated from the glucose concentration with gaussian white noise. AC_Mean and AC_Var calculated in each trial are shown in the left panels. The distributions of AC_Mean and AC_Var are shown in the right. The blue is simulated using the parameters reported as the average values for healthy subjects. The green is simulated at half the average values. The *P* values are for testing the hypothesis of no difference between the two groups.

We also investigated the effects of noise on AC_Mean and AC_Var (Supplementary text 3, Figs. 6C, S13 and S14), and found that AC_Var was more robust to noise than AC_Mean, consistent with the result that AC_Var exhibited a stronger correlation with DI than AC_Mean (Fig. 2A). Moreover, extending the measurement period and decreasing the measurement interval of CGM were found to enhance the accuracy of reduced blood glucose control ability (Supplementary text 3 and Fig. S14).

### Web application for calculating CGM-derived indices

To easily calculate CGM-derived indices, we developed a web application (https://cgm-ac-mean-std.streamlit.app/) that calculates CGM_Mean, CGM_Std, AC_Mean, and AC_Var (Fig. S15). This application was implemented in streamlit. In using this application, glucose should be measured every 5 min. The application can also run on a local machine using the code in GitHub repository (https://github.com/HikaruSugimoto/CGM_AC).

## DISCUSSION

In this study, we found that CGM-derived indices, including AC_Mean and AC_Var, were effective in identifying individuals with reduced blood glucose control ability within a population predominantly consisting of individuals with NGT. AC_Mean exhibited a significant correlation with insulin clearance, and AC_Var was significantly correlated with both insulin clearance and DI. Insulin clearance predicts the incidence of T2DM in non-DM subjects,^36^ and DI predicts the development of future T2DM beyond FBG and PG120.^37^ Collectively, these findings suggest that CGM-derived indices, including AC_Mean and AC_Var, can identify abnormalities in blood glucose regulation at an early stage, potentially serving as alternatives to single-point blood tests or OGTT, which have been found to be inadequate^9–11^ or inconvenient^12^ for the screening of pre-DM.

We also predicted DI and glucose dysregulation by using multiple linear regression, PLS regression, Lasso regression, random forests, and logistic regression with L1 regularization. DI is suggested to be a predictor of T2DM development,^37^ and decreases in insulin sensitivity and insulin secretion reportedly precede the onset of T2DM.^1^ However, accurate measurement of DI, insulin sensitivity, and insulin secretion are laborious. The prediction models in this study included only indices derived from a single-point blood test, physical examinations, and CGM, which are relatively easy-to-measure variables. Given that the accuracy of the linear regression model with CGM_Std and AC_Var as input variables and clamp DI as the objective variable was about the same as the accuracy of predicting clamp DI from oral DI, these relatively easy-to-measure variables may be alternatives to OGTT and clamp tests under some conditions. In this study, we investigated two different datasets, but both datasets were small in size and included only three T2DM subjects (Table S1). The main purpose of this study was not to create a predictor that can be used in clinical settings; rather, this exploratory study showed that reduced blood glucose control ability may be predictable using CGM and that AC_Mean and AC_Var can be useful in the prediction. Larger studies are required for more accurate predictions of the abnormalities.

AC_Mean and AC_Var exhibited low collinearity with other CGM-derived indices and showed the potential to capture changes in blood glucose dynamics indicative of early-stage glucose intolerance (Figs. 2–5). The mathematical modeling further highlighted their potential in identifying abnormalities even when FBG levels remain unchanged (Fig. 6). Collectively, these results indicate that AC_Mean and AC_Var can capture glycemic disability that cannot be captured by the conventional indices. However, the exact underlying mechanisms of these results and their relationship with other CGM-derived indices remain unknown. In this study, we investigated only the mean and the variance of the autocorrelation function at lags 1–30, relatively well-known CGM-derived indices,^25^ and three indices that have been reported to identify abnormal glucose regulation.^19^ However, the mean and the variance of the autocorrelation function at lags 1–2 (Fig. S4) were also significantly correlated with oral DI (r = –0.29; *P* = 0.02 and r = 0.29, *P* = 0.02, respectively), and other CGM-derived indices of glycemic variability have been reported.^26,45–56^ It is necessary to investigate these indices comprehensively to determine the extent to which abnormalities can be identified from CGM in the future.

In conclusion, the current study demonstrated that CGM-derived indices, AC_Mean and AC_Var, can detect decreased blood glucose control ability beyond conventional markers such as FBG, HbA1c, and other CGM-derived indices. Furthermore, the results indicate that CGM could potentially estimate DI, a strong indicator of glucose dysregulation and T2DM onset. This suggests that CGM-derived DI (CGM DI) may serve as an alternative to the labor-intensive measurements involved in conventional DI assessment using OGTT (oral DI) or clamp tests (clamp DI).

## Data Availability

The CGM data that support the findings of this study are available from the GitHub repository (https://github.com/HikaruSugimoto/CGM_AC). The code that calculates AC_Mean and AC_Var is also available from the repository (https://github.com/HikaruSugimoto/CGM_AC) and the web application (https://cgm-ac-mean-std.streamlit.app/).

## Acknowledgments

**Personal Thanks.** We thank Mio Shudo and Yuka Nakamura for their assistance with the analysis; and our laboratory members for critically reading this manuscript

## Author Contributions

H.S. analyzed the data. H.S., K.H., T.N., T.Y., H.M., N.O-S., M.F., Y.H., K.S., W.O., and S.K. wrote the manuscript. T.N., T.Y., H.M, and N.O-S. conducted clinical examinations. W.O. and S.K. supervised the study.

## Funding and Assistance

This study was supported by the Japan Society for the Promotion of Science (JSPS) KAKENHI (JP21H04759), CREST, the Japan Science and Technology Agency (JST) (JPMJCR2123), and The Uehara Memorial Foundation.

