## Supplementary file for "Improved Detection of Decreased Glucose Handling Capacities via Novel Continuous Glucose Monitoring-Derived Indices: AC_Mean and AC_Var"

#### Supplementary Texts

##### 1. Detailed procedures for a 75-g oral glucose tolerance test (OGTT), a consecutive hyperglycemic and hyperinsulinemic-euglycemic clamp, and a continuous glucose monitoring (CGM)

In a standard 75-g OGTT, blood samples were collected at 30, 60, 90, and 120 minutes after glucose ingestion for measuring the levels of plasma glucose, serum insulin, and C-peptide reactive protein (CPR) (measured hexokinase UV method (Kanto Chemical Industry Co., Ltd., Japan), chemiluminescent enzyme immunoassay (Lumipulse Presto insulin, Fujirebio, Japan), chemiluminescent enzyme immunoassay (Lumipulse Presto insulin, Fujirebio, Japan), respectively).

The hyperglycemic and hyperinsulinemic-euglycemic clamp analysis was performed with the use of an artificial endocrine pancreas (STG-55; Nikkiso Co., Ltd, Tokyo, Japan) as described previously[1]. In brief, from 0 to 90 min, a hyperglycemic clamp was performed by intravenous infusion of a bolus of glucose (9622 $mg/m^2$) within 15 min followed by that of a variable amount of glucose to maintain the plasma glucose level at 200 mg/dL. Ten minutes after the end of the hyperglycemic clamp, a 120-min hyperinsulinemic-euglycemic clamp was initiated by intravenous infusion of human regular insulin (Humulin R, Eli Lilly Japan K.K., Kobe, Japan) at a rate of 40 mU $m^{-2}$ $min^{-1}$ and the hyperinsulinemic state was maintained to achieve a target glucose level of the fasting level and a serum insulin concentration of 100 μU/ml. Plasma glucose concentrations were measured every minute during the clamp and averaged over a 5-minute period. The data on plasma glucose, serum insulin, and CPR were also collected before and at 5, 10, 15, 60, 75, 90, 100, 190, and 220 minutes after the onset of the clamp tests.

Data from the CGM were used in the analysis for the 72-hour period from the next days fitted with iPro. At least three times a day, capillary blood glucose levels were measured using a glucometer (Accucheck Performa, Roche Diabetes Care Japan K.K., Tokyo, Japan), which was necessary to calibrate the CGM system.

**2. Prediction of glycemic disability using random forests or a logistic regression analysis with L1 regularization**

To predict glycemic disability and to estimate the non-linear relationship among the variables shown in Figure 3A in predicting glycemic disability, we conducted random forests, which have been used to predict T2DM and its complications, and to estimate risk factors associated with T2DM.[2–4] Decrease in insulin secretion and insulin sensitivity are reported to start years before diabetes development and already present in the pre-DM stage,[5] and in this study, some subjects diagnosed with NGT had relatively low I.I. and composite index (Fig. S8A). Hence, in this study, we defined glycemic disability as follows; I.I. < 0.4, composite index < 3.0, FBG > 110 mg/dL, or PG120 > 140 mg/dL (Fig. S8A and Fig. S8B). Of note, there are no set standards for glycemic disability, but these values are within the range of values defined as abnormal in previous studies.[6–8]

The maximum leaf nodes of the 6 and the features with AC_Mean and AC_Var provided the relatively better performance, with the accuracy and the F1 score were 0.75 and 0.51, respectively (Fig. S9A). To investigate the importance of each feature in estimating the anomaly, we calculated the permutation importance and the feature importance (Fig. S9B). For both indices, a higher value indicates that the feature is more important in the prediction. The permutation importance and the feature importance of AC_Mean in predicting the glycemic disability were relatively high (Fig. S9B). The permutation importance of AC_Mean was higher than FBG, which is included in the objective function. To further investigate the importance of AC_Mean and AC_Var in the prediction, we also conducted Boruta, an algorithm for finding features carrying information usable for the prediction.[9] In this analysis, the maximum of the shadow feature was used as a threshold in deciding whether the input variables were better at prediction than the shadow ones, which could be a harsh threshold;[9] nevertheless, AC_Mean was statistically significantly contributed to the prediction (Fig. S9B).

To validate the importance of AC_Mean and AC_Var in estimating glycemic disability, we also conducted a logistic regression analysis with L1 regularization (Fig. S10). The leave-one-out cross validation indicated that the optimal regularization coefficients, lambda, was 18.5 (Fig. S10A, dashed line). At this lambda, the coefficient of AC_Var was estimated as non-zero coefficient (Fig. S10B, C). Collectively, these results suggest that examining these two variables in addition to conventional indices may predict the anomalies more accurately.

**3. Effect of noise on AC_Mean and AC_Var**

We investigated the effect of noise on AC_Mean and AC_Var. AC_Mean and AC_Var of blood glucose dynamics simulated using the mean values of $k_{\mathrm{sec}}k_{\mathrm{sen}}$ and $k_{\mathrm{cle}}$ at NGT were compared with those simulated by using the half values of the parameters. Applying gaussian white noise with the mean of 0 and the variance of 1 (Fig. S13A), followed by the calculation of AC_Mean and AC_Var was repeated 1000 times. Since the exact modeling of the statistical properties of CGM sensor errors is difficult,[10] we only investigated the properties of AC_Mean and AC_Var at different white noise magnitudes. Of note, the blood glucose difference due to the differences in $k_{\mathrm{sec}}k_{\mathrm{sen}}$ and $k_{\mathrm{cle}}$ was at most 0.45 mg/dL, and the variance of the difference was only 0.014. Even if the applied noise is greater than the difference between the two groups (Fig. S13B), AC_Var in particular could distinguish the two groups (Fig. 6C, $P < 0.01$). Cohen's d, which has been used to estimate the effect size, was larger in AC_Var (Fig. 6C). As the variance of the noise was reduced, the Cohen's d of both AC_Mean and AC_Var became larger (Fig. S14A-D). As the measurement period was increased from 24-hour to 72-hour (Fig. S14E), or as the measurement interval was reduced from every 5 minutes to every 1 minute (Fig. S14F), the Cohen's d of both AC_Mean and AC_Var became larger. Under all conditions (Figs. 6, S14), the Cohen's d of AC_Var was larger than that of AC_Mean, suggesting that AC_Var is more robust to noise than AC_Mean. This is consistent with the result that AC_Var was more correlated with DI (Fig. 2A).

#### Supplementary Figures

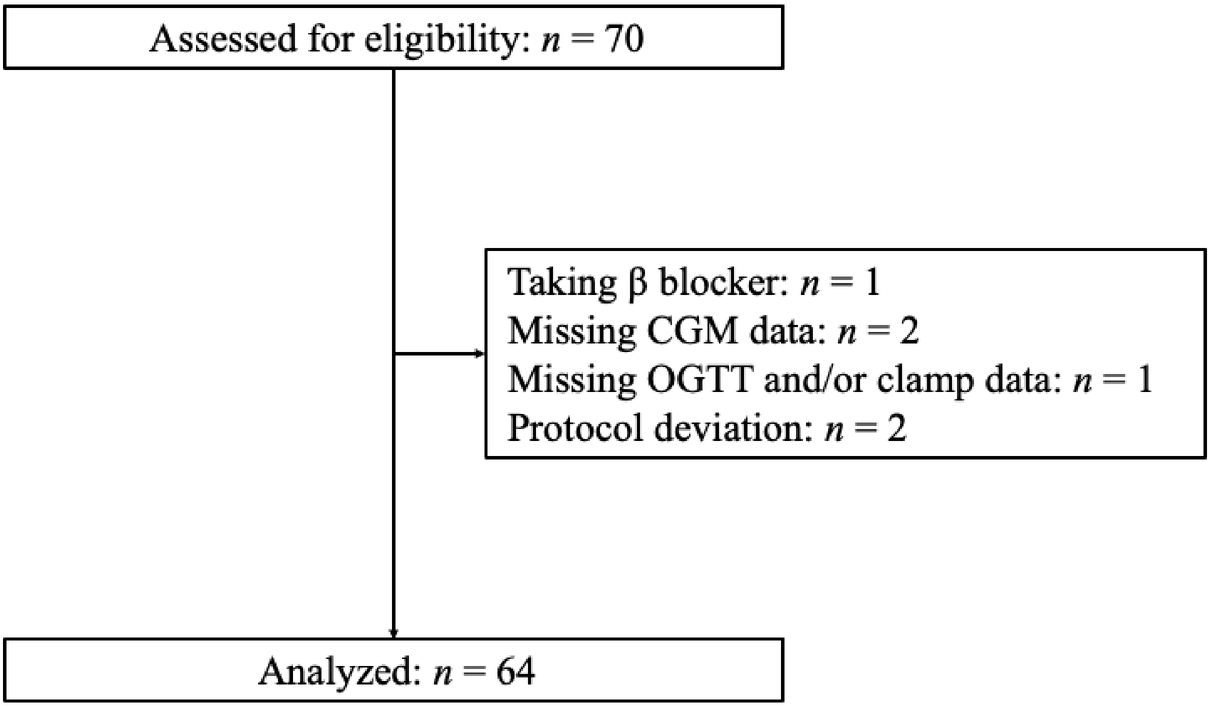

**Figure S1. Flow diagram of participant recruitment.**

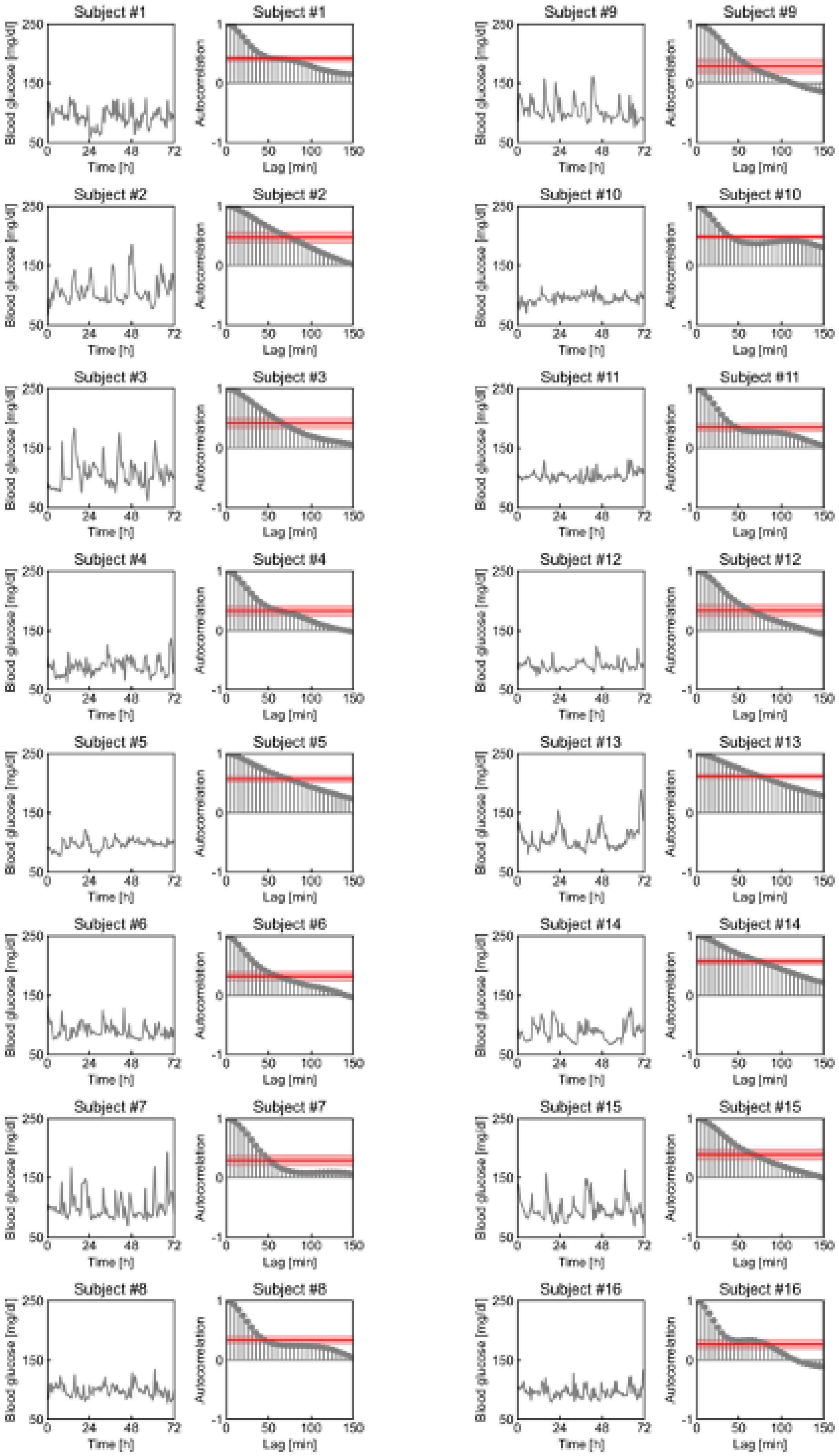

Subject #1
Subject #2
Subject #3
Subject #4
Subject #5
Subject #6
Subject #7
Subject #8
Subject #9
Subject #10
Subject #11
Subject #12
Subject #13
Subject #14
Subject #15
Subject #16
Blood glucose [mg/dl]
Time [h]
Autocorrelation
Lag [min]

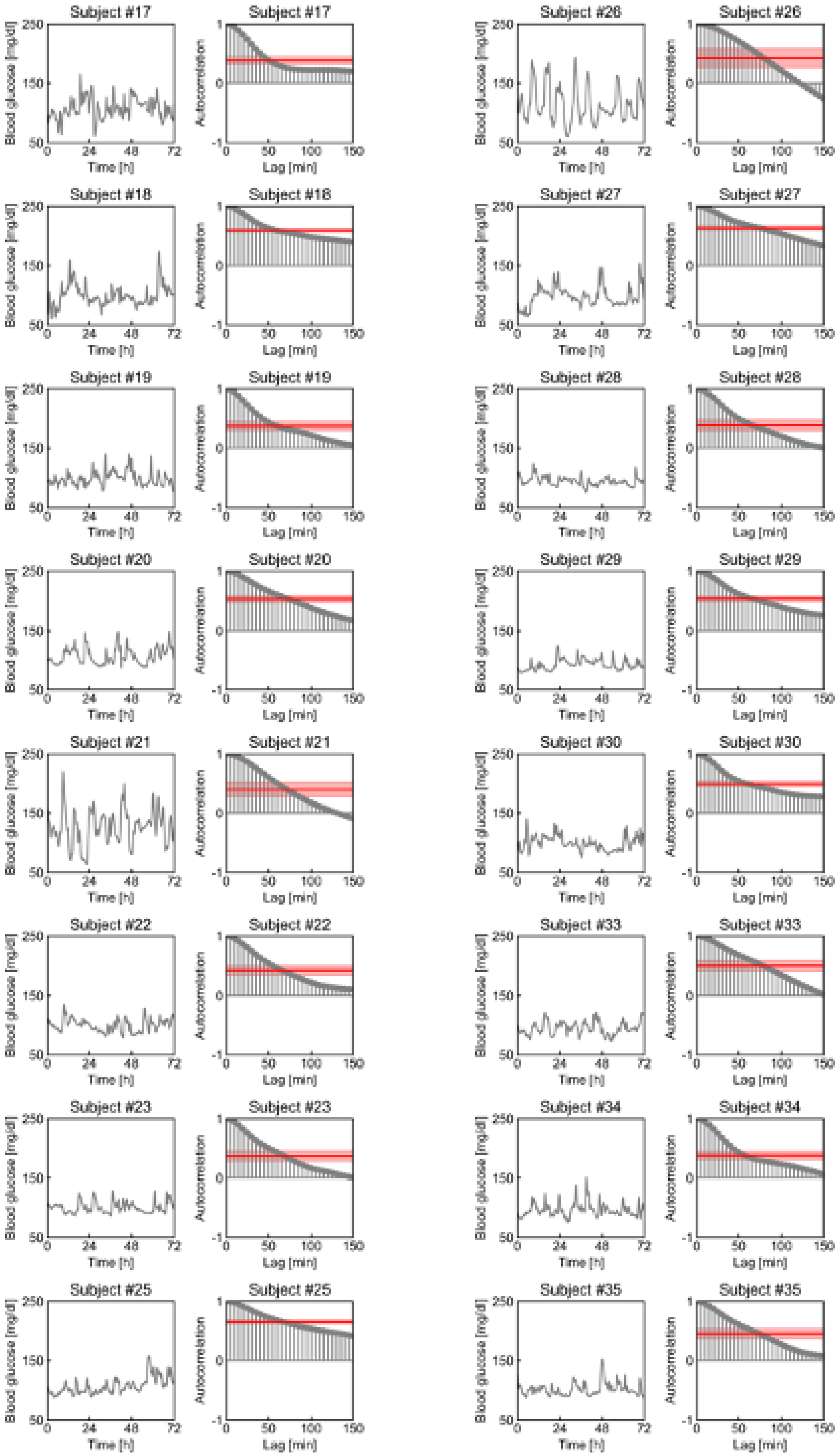

Subject #17
Subject #17
Subject #26
Subject #26
Subject #18
Subject #18
Subject #27
Subject #27
Subject #19
Subject #19
Subject #28
Subject #28
Subject #20
Subject #20
Subject #29
Subject #29
Subject #21
Subject #21
Subject #30
Subject #30
Subject #22
Subject #22
Subject #33
Subject #33
Subject #23
Subject #23
Subject #34
Subject #34
Subject #25
Subject #25
Subject #35
Subject #35
Blood glucose [mg/dl]
Time [h]
Autocorrelation
Lag [min]

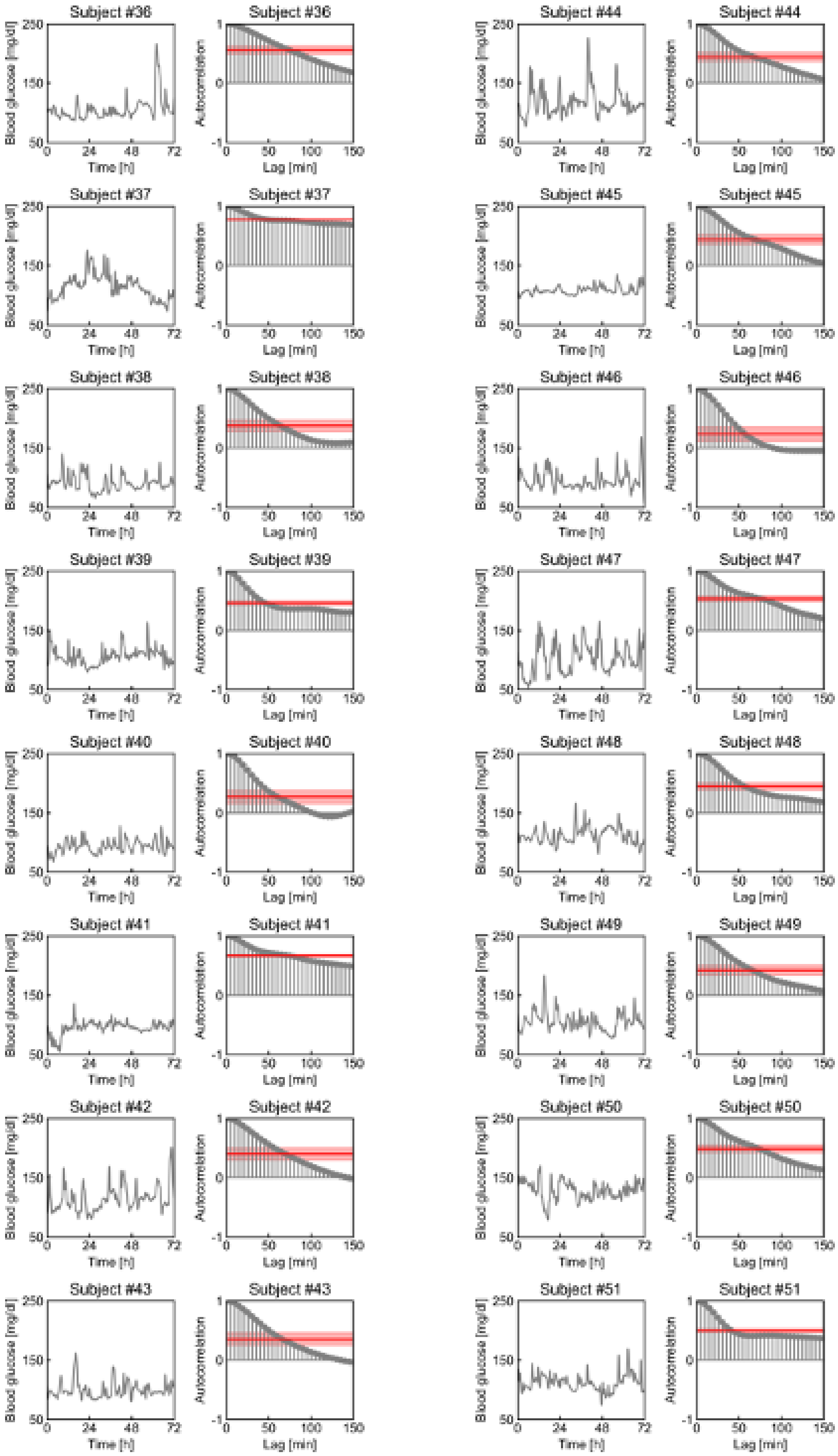
Subject #36
Subject #37
Subject #38
Subject #39
Subject #40
Subject #41
Subject #42
Subject #43
Subject #44
Subject #45
Subject #46
Subject #47
Subject #48
Subject #49
Subject #50
Subject #51
Blood glucose [mg/dl]
Time [h]
Autocorrelation
Lag [min]

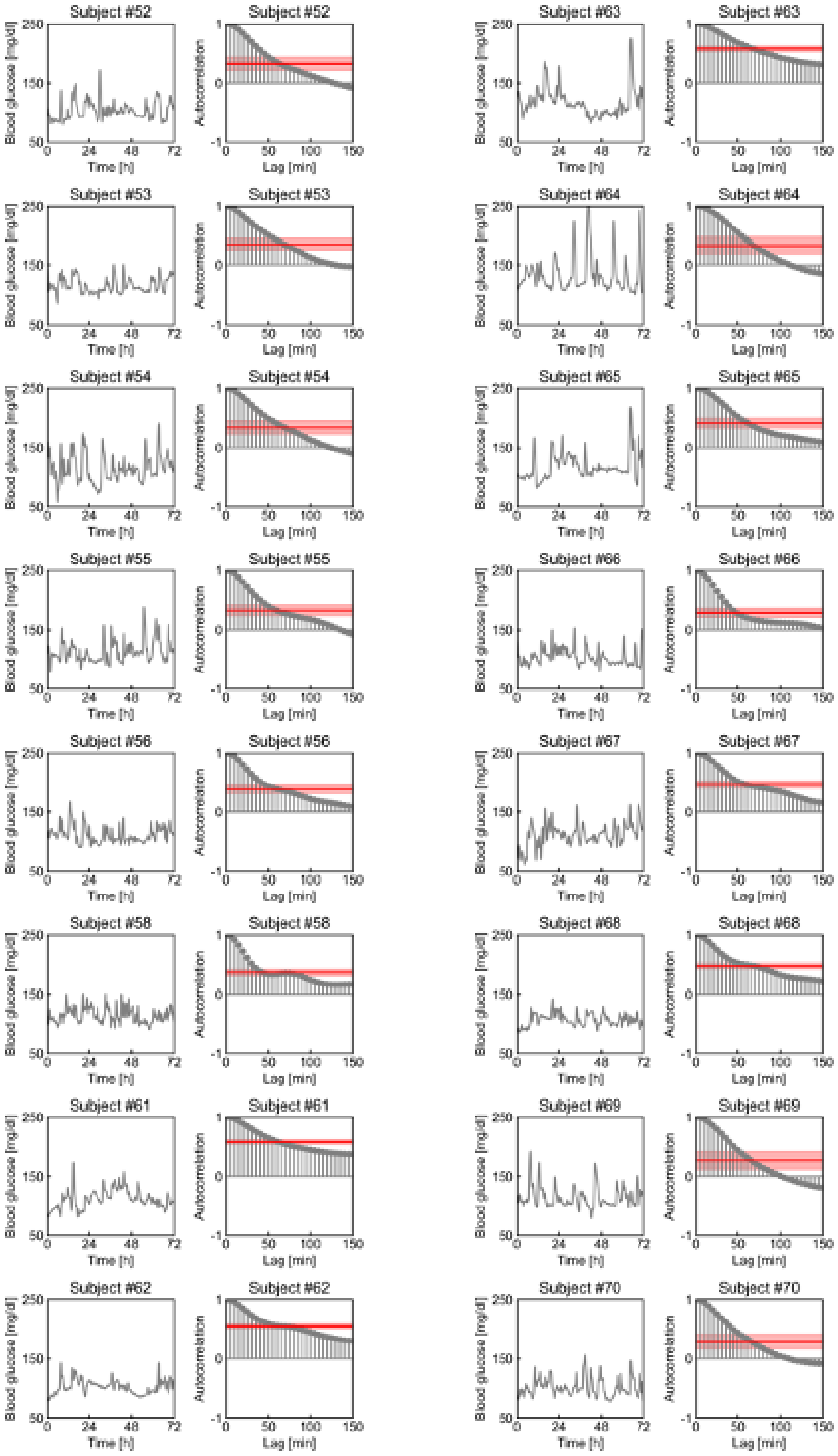
Subject #52
Subject #53
Subject #54
Subject #55
Subject #56
Subject #58
Subject #61
Subject #62
Subject #63
Subject #64
Subject #65
Subject #66
Subject #67
Subject #68
Subject #69
Subject #70
Blood glucose [mg/dl]
Time [h]
Autocorrelation
Lag [min]

**Figure S2. Blood glucose fluctuations and correlogram of the time series data for each subject.**

Time courses of blood glucose levels measured by CGM and their correlograms. Red lines indicate the mean value of the autocorrelation functions (AC_Mean), and red shaded areas indicate the variance value of the autocorrelation functions (AC_Var).

109

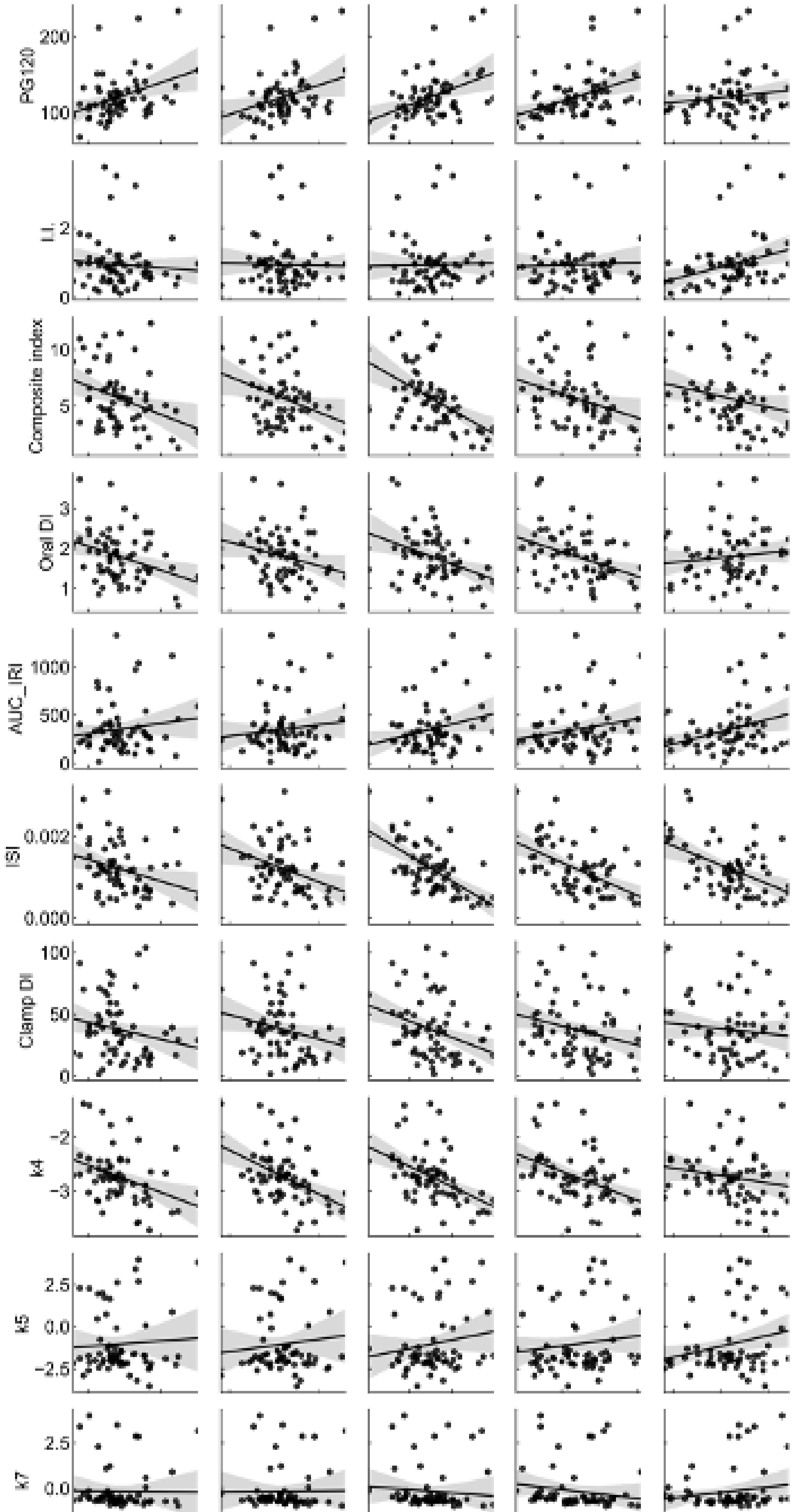

PG120
LI
Composite index
Oral DI
AUC_IRI
ISI
Clamp DI
k4
k5
k7
SBP
DBP
BMI
Acir
Body fat percentage

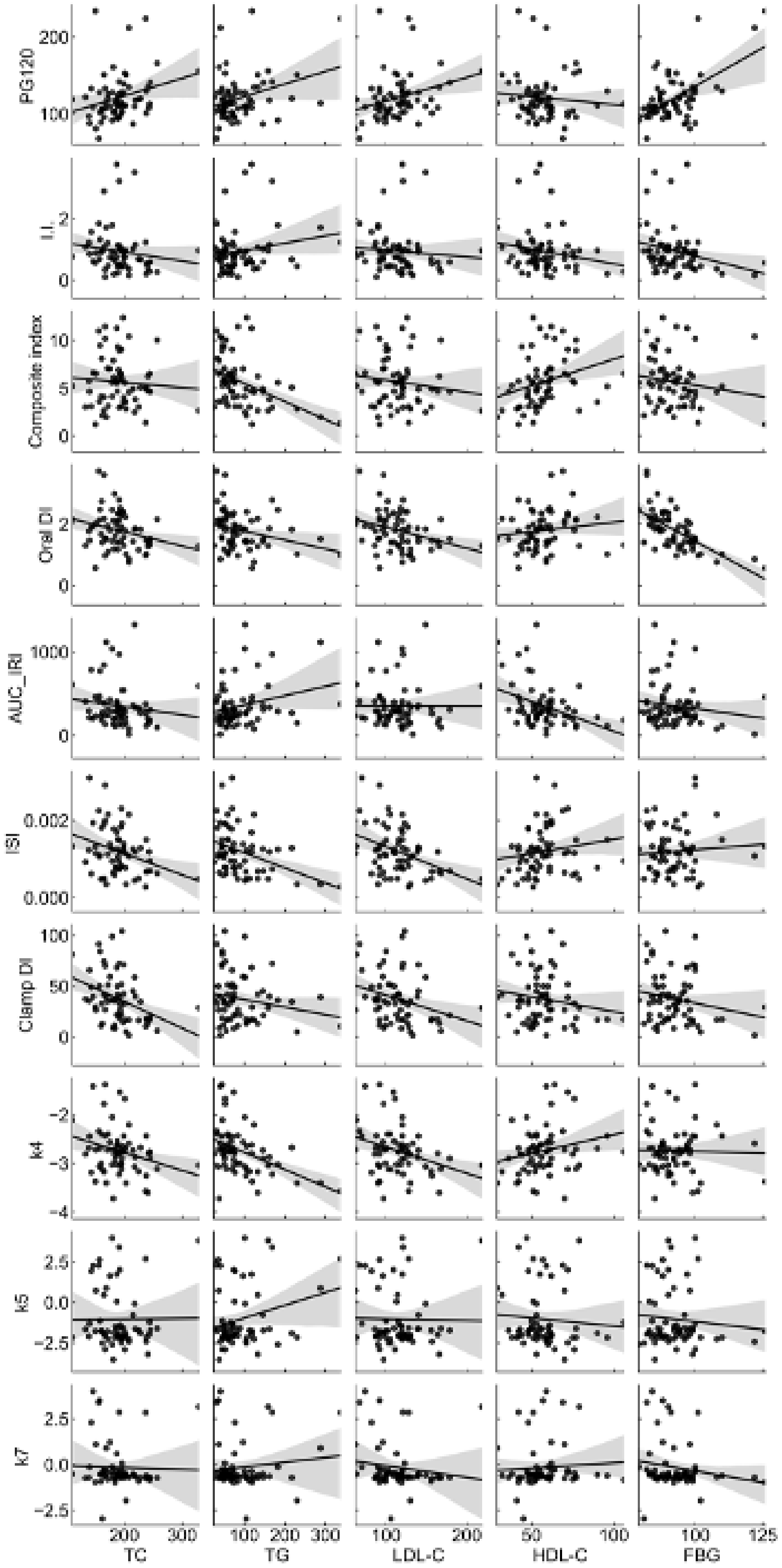

PG120
I.I.
Composite index
Oral DI
AUC_IRI
ISI
Clamp DI
k4
k5
k7
TC
TG
LDL-C
HDL-C
FBG

110

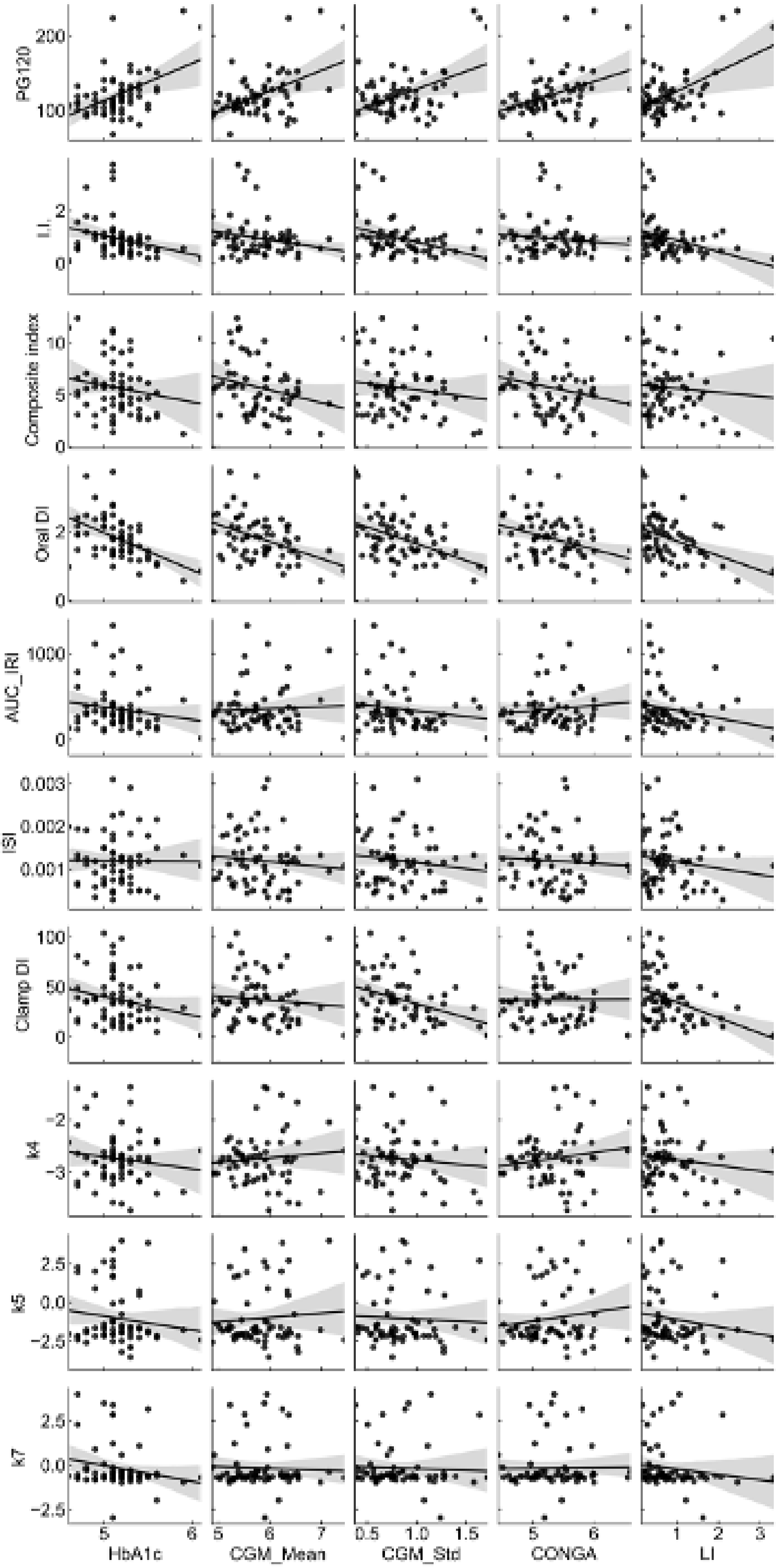
PG120
I.I.
Composite index
Oral DI
AUC_IRI
ISI
Clamp DI
k4
k5
k7
HbA1c
CGM_Mean
CGM_Std
CONGA
LI

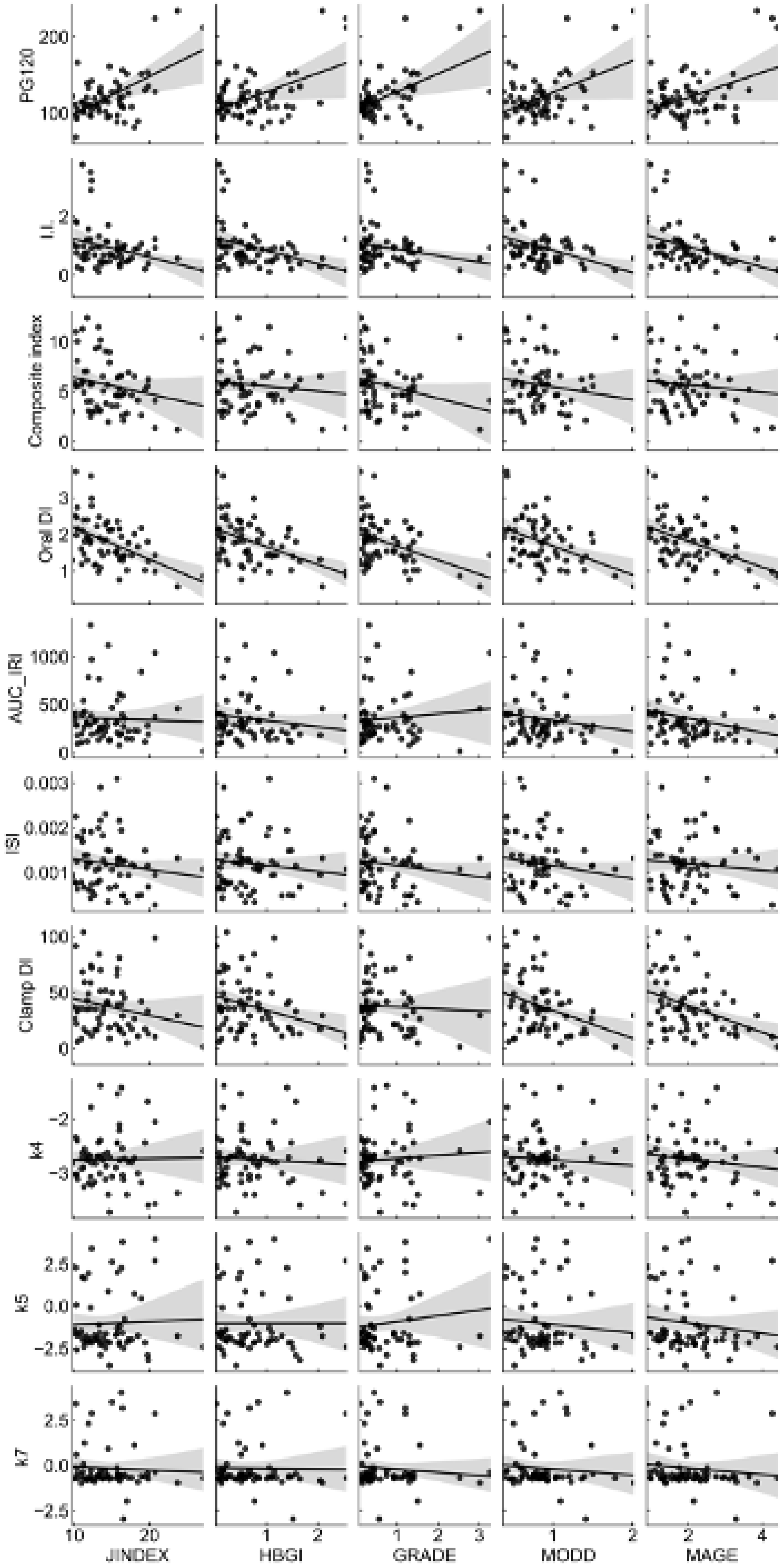
PG120
I.I.
Composite index
Oral DI
AUC_IRI
ISI
Clamp DI
k4
k5
k7
JINDEX
HBGI
GRADE
MODD
MAGE

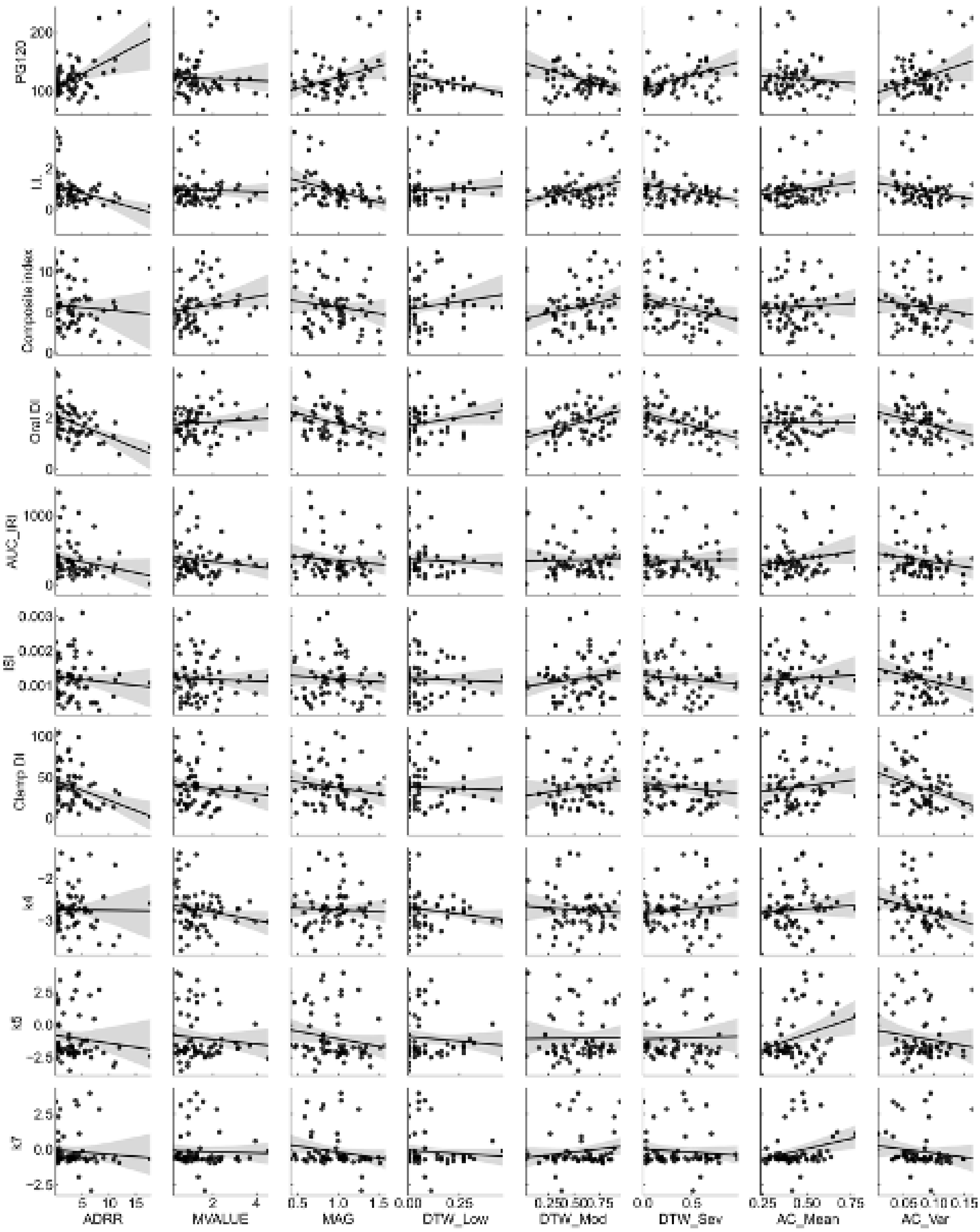

**Figure S3. Relation between the indices from CGM and those from OGTT and clamp tests.**

Scatter plots and fitted linear regression lines for CGM-derived, OGTT-derived, and clamp tests-derived indices. Each point corresponds to the values for a single subject. Gray shaded area indicates 95% confidential interval.

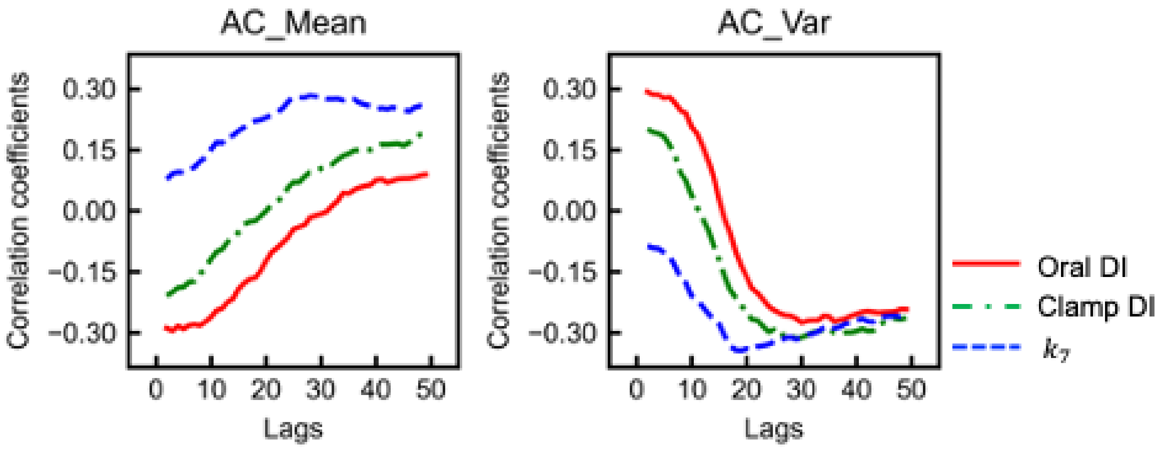

**Figure S4. Characteristics of AC_Mean and AC_Var when the lags are different.**

The effect of lags on the Spearman's correlation coefficient between AC_Mean (the left) or AC_Var (the right) and oral DI, clamp DI, or $k_7$. Oral DI, oral disposition index; clamp DI, clamp disposition index. $k_7$ corresponds to insulin clearance.

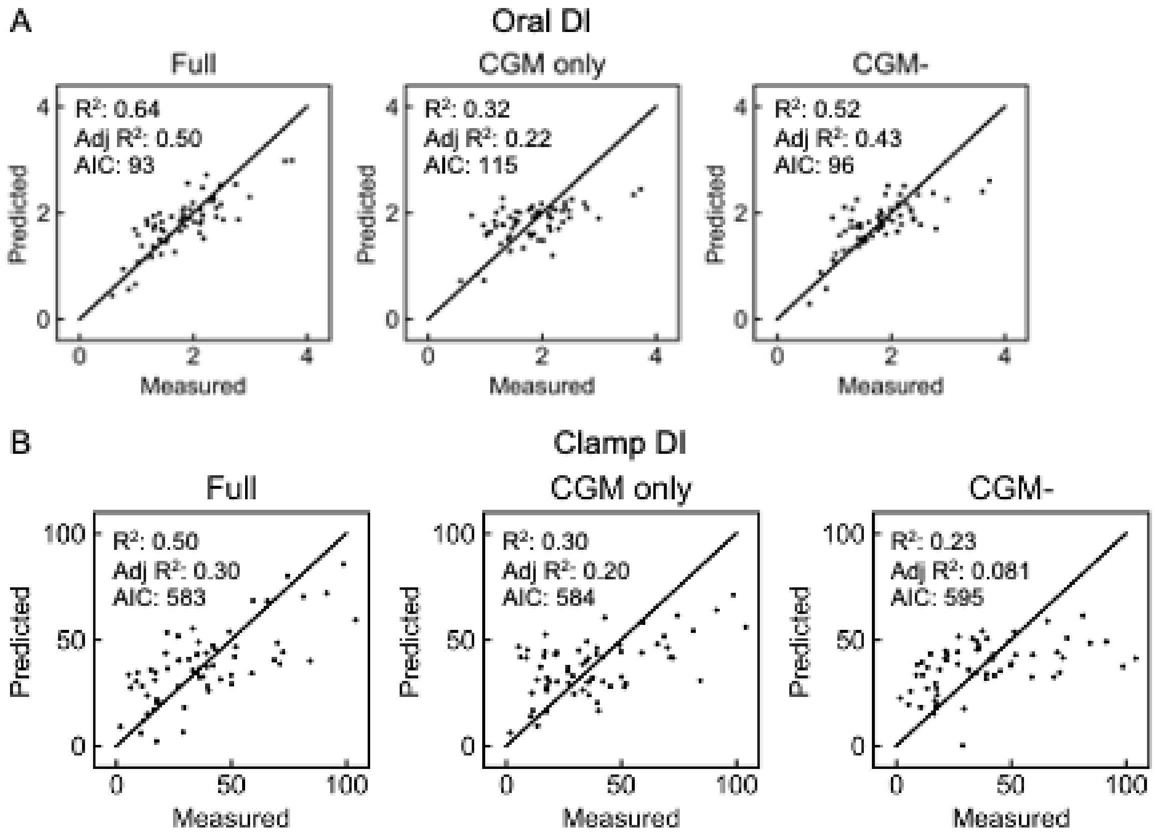

**Figure S5. Multiple linear regression analyses for predicting disposition index from a single blood test, physical measurements, and CGM-derived indices.**

(A) Scatter plots for predicted oral DI versus measured oral DI. Each point corresponds to the values for a single subject. The left (Full) was predicted using the 18 variables, including a single blood test, physical measurement, and CGM-derived indices, as shown in Fig. 3B. The middle (CGM only) was predicted using the indices from CGM (Fig. S6B), and the right (CGM-) was predicted using the indices from a single blood test and physical measurements (Fig. S6D). (B) Scatter plots for predicted clamp DI versus measured clamp DI. $R^2$, the coefficient of determination; Adj $R^2$, the adjusted coefficient of determination; AIC, Akaike Information Criteria; oral DI, oral disposition index; clamp DI, clamp disposition index.

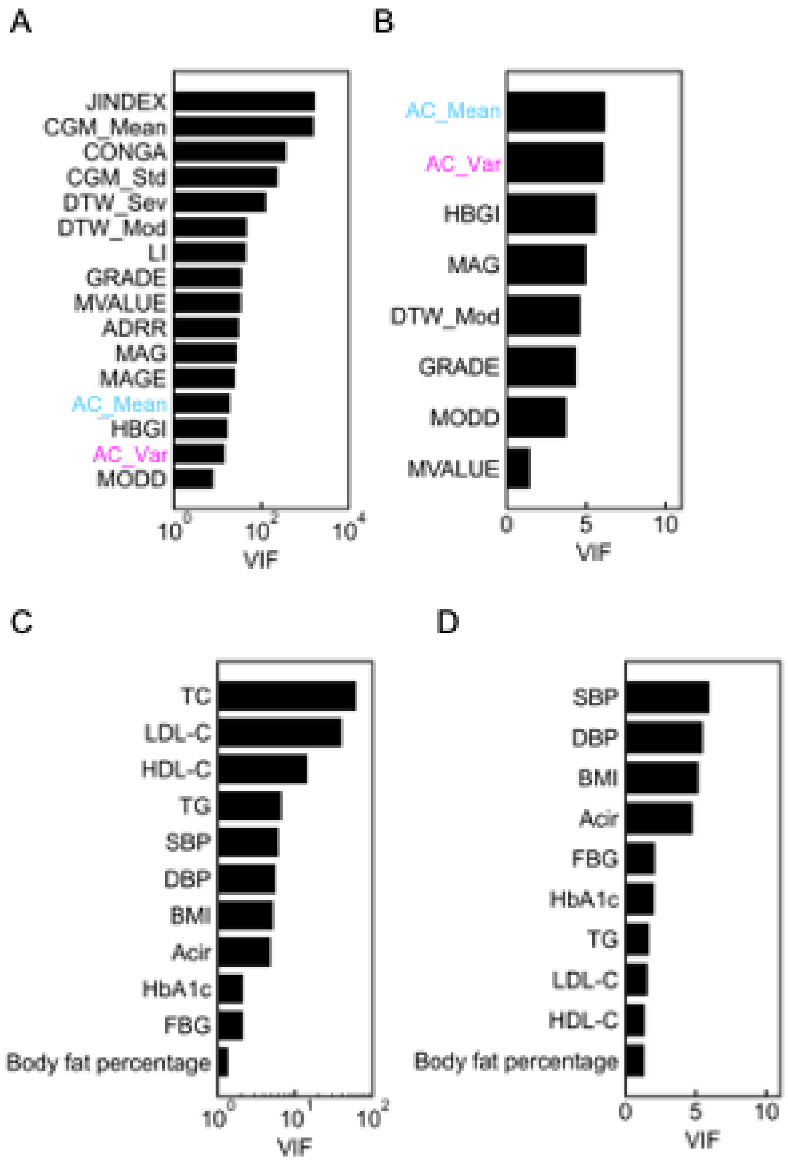

**Figure S6. Relation among indices from a single blood test, physical measurements, and CGM-derived.**

Variance inflation factor (VIF) of CGM-derived indices (A), and that of each variable remaining after removing the variable with the highest VIF one by one until the VIF of all variables are less than 10 (B). VIF of indices from a single blood test and physical measurement (C), and that of each variable remaining after removing the variable with the highest VIF one by one until the VIF of all variables are less than 10 (D).

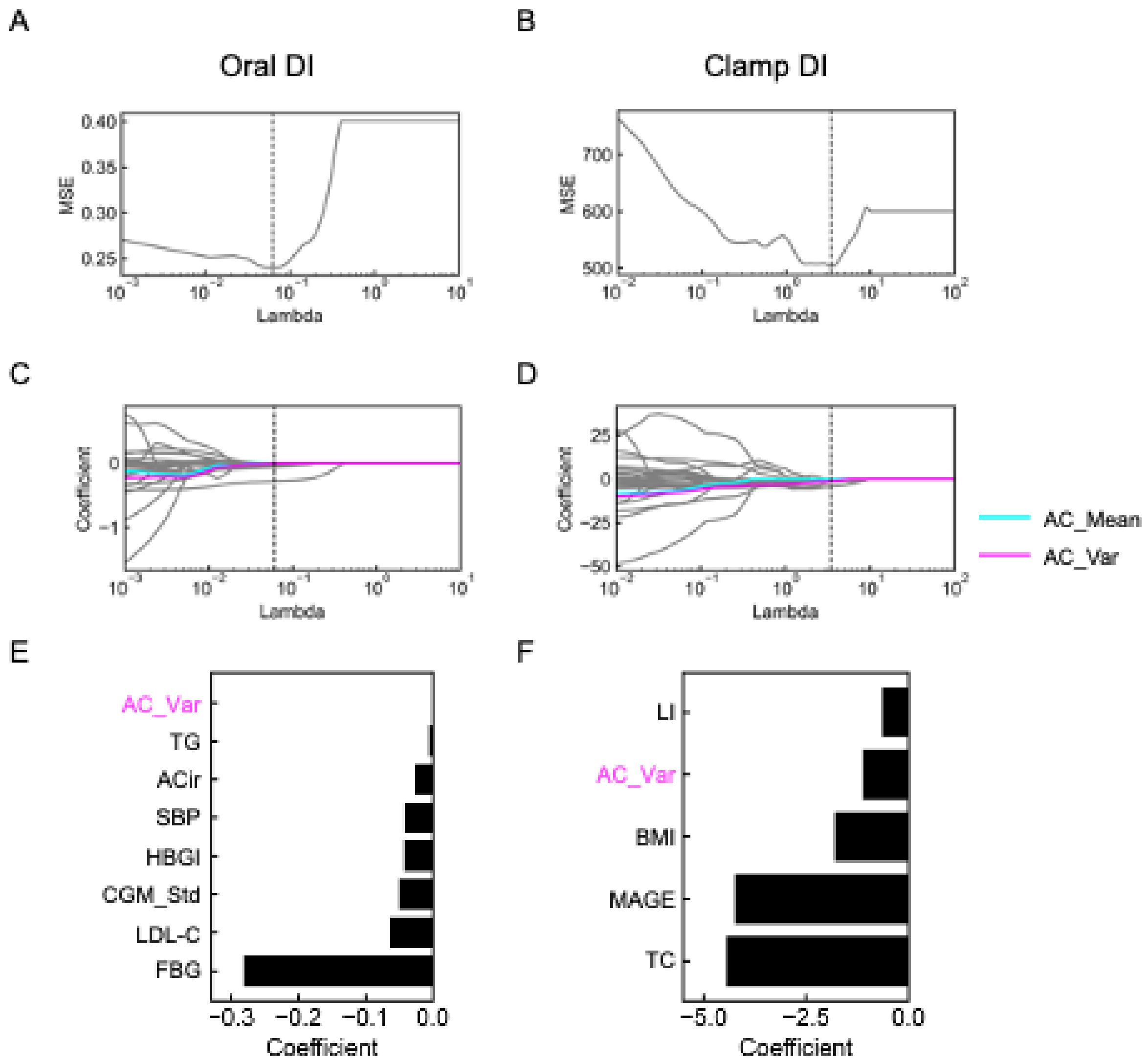

**Figure S7. Lasso regression analyses for predicting disposition index.**

Relation between regularization coefficients (Lambda) and the mean squared error (MSE) based on the leave-one-out cross validation in predicting oral DI (A) and clamp DI (B). Dotted vertical lines indicate the optimal lambda which provides the least MSE. The optimal lambda for oral DI was 0.061 (A) and that for clamp DI was 3.49 (B). Lasso regularization paths along the Lambda in predicting oral DI (C) and clamp DI (D). Cyan, magenta, and gray lines indicate the estimated coefficients of AC_Mean, AC_Var, and the other input variables, respectively. Dotted vertical lines indicate the optimal Lambda. Estimated coefficients with the optimal Lambda in predicting oral DI (E) and clamp DI (F). Only variables with non-zero coefficients are shown.

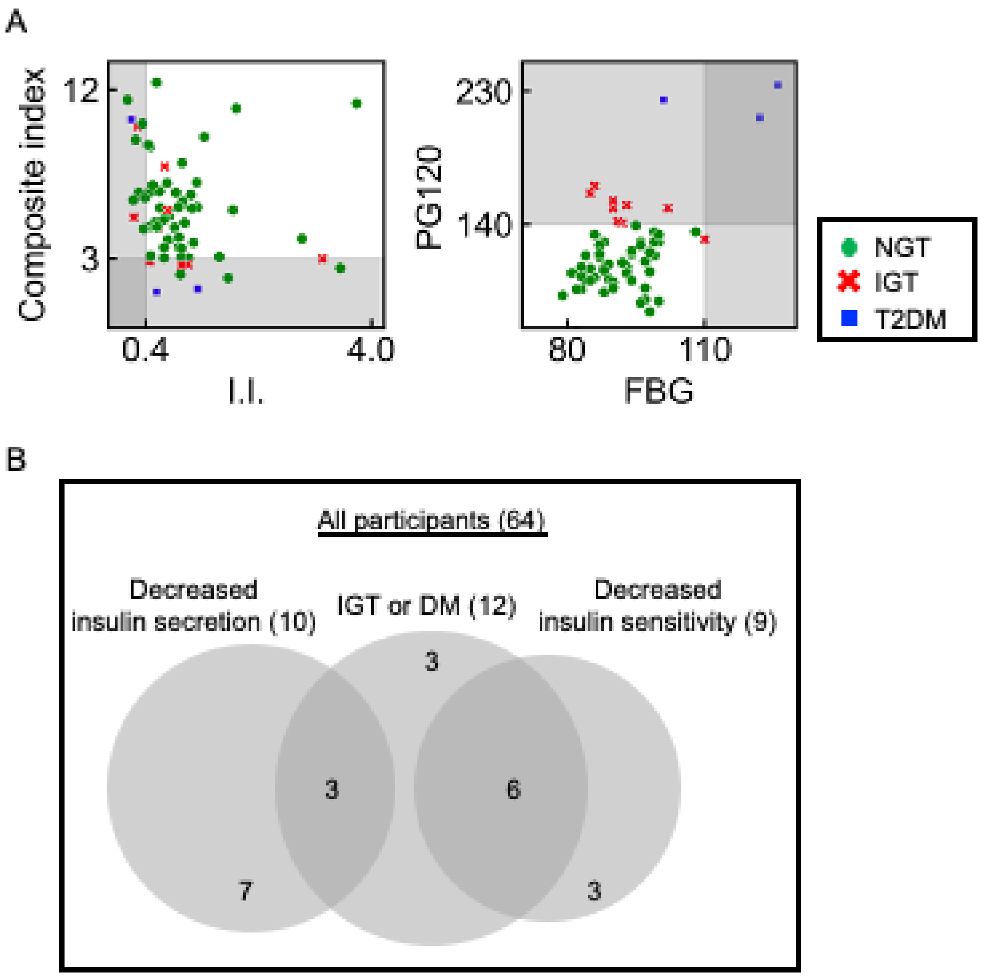

**Figure S8. Insulin secretion, insulin sensitivity, and blood glucose levels of the subjects in this study.**

(A) Scatter plots for insulinogenic index (I.I.) versus composite index (the left), and fasting blood glucose (FBG) versus plasma glucose concentration at 120 min during the OGTT (PG120) (the right). Each point corresponds to the values for a single subject. Green circles, red crosses, and blue squares indicate NGT, IGT, and T2DM subjects, respectively. Gray shaded areas indicate the range of values for glycemic disability defined in this study (I.I. < 0.4, composite index < 3.0, FBG > 110 mg/dL, or PG120 > 140 mg/dL). (B) Venn diagram indicating the number of subjects with reduced insulin secretion (I.I. < 0.4), reduced insulin sensitivity (composite index < 3.0), IGT, or T2DM (FBG > 110 mg/dL or PG120 > 140 mg/dL).

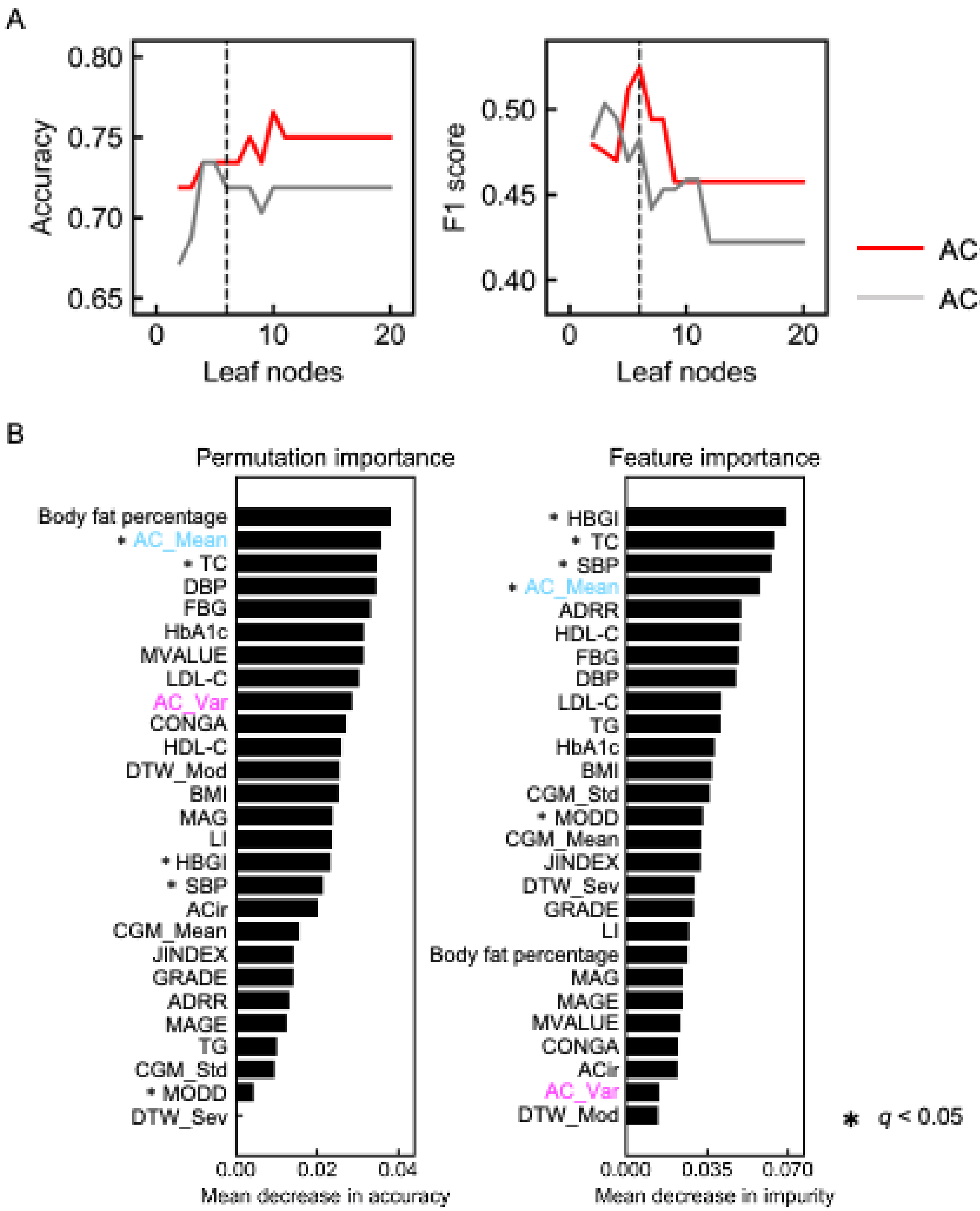

**Figure S9. Random forests classification for glycemic dysregulation.**

(A) Effect of leaf nodes number and the features, AC_Mean and AC_Var, on the performance of random forests classification. Accuracy and F1 score are based on leave-one-out cross-validation. (B) Importance of the input variables in the random forests model. X-axis represents the permutation importance (left panel) and the feature importance (right panel). Y-axis represents the variables. The permutation importance reflects decreases in classification performance when the values of a given variable have been randomly permuted. Feature importance was based on the Gini index. FBG, fasting blood glucose; ACir, abdomen circumference; BMI, body mass index; SBP, systolic blood pressure; DBP, diastolic blood pressure; TC, total cholesterol; TG, triglycerides; LDL-C, low-density lipoprotein cholesterol; HDL-C, high-density lipoprotein cholesterol. The $q$ values, which are calculated by Boruta, are for testing the hypothesis of no relevance to glycemic dysregulation.

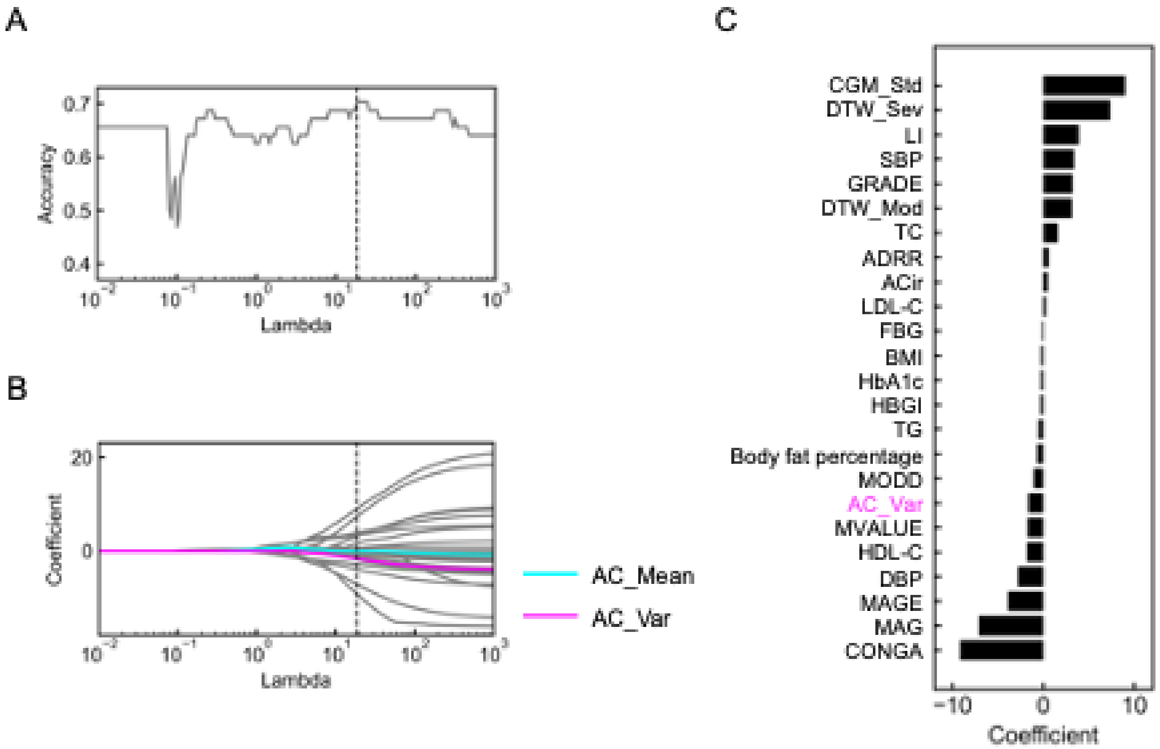

**Figure S10. Logistic regression analyses with L1 regularization for predicting glycemic disability.**

(A) Relation between regularization coefficients (Lambda) and the accuracy in predicting glycemic disability based on the leave-one-out cross validation. Dotted vertical lines indicate the optimal lambda which provides the best accuracy. (B) Regularization paths along the lambda in predicting glycemic disability. Cyan, magenta, and gray lines indicate the estimated coefficients of AC_Mean, AC_Var, and the other input variables, respectively. Dotted vertical line indicates the optimal lambda. (C) Estimated coefficients with the optimal lambda in predicting glycemic disability. Only variables with non-zero coefficients were shown.

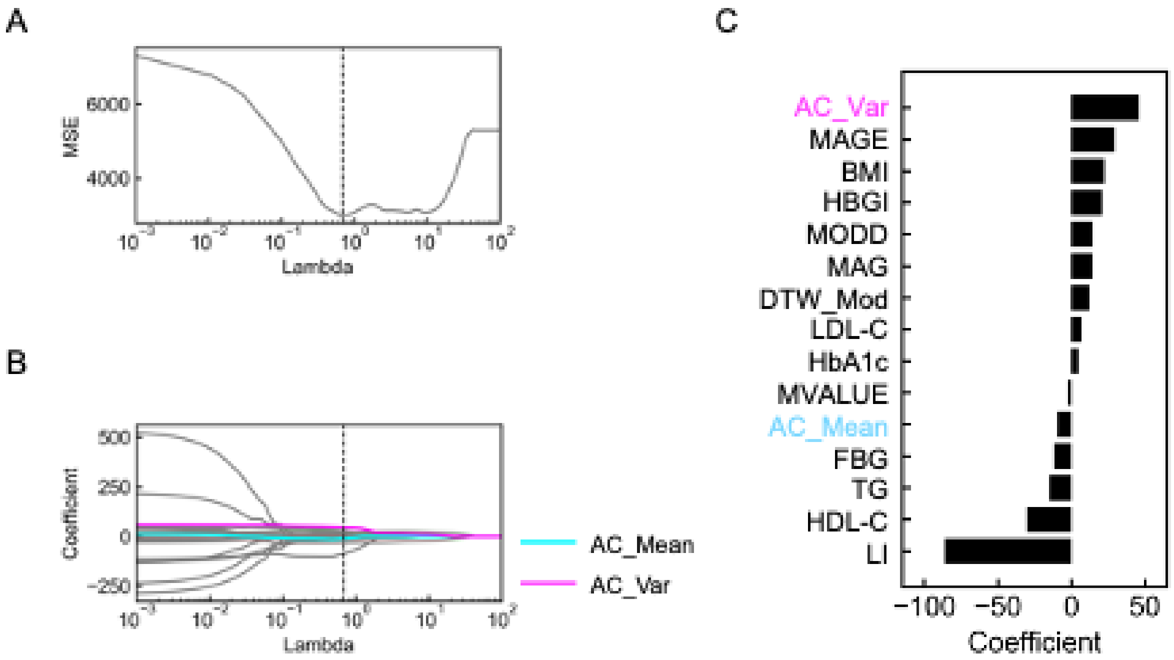

**Figure S11. Lasso regression analyses for predicting SSPG.**

(A) Relation between regularization coefficients (Lambda) and the mean squared error (MSE) based on the leave-one-out cross validation in predicting SSPG. Dotted vertical line indicates the optimal Lambda which provides the least MSE. The optimal Lambda for SSPG was 0.69. (B) Lasso regularization path along the Lambda in predicting SSPG. Cyan, magenta, and gray lines indicate the estimated coefficients of AC_Mean, AC_Var, and the other input variables, respectively. Dotted vertical line indicates the optimal Lambda. (C) Estimated coefficients with the optimal Lambda in predicting SSPG. Only variables with non-zero coefficients are shown.

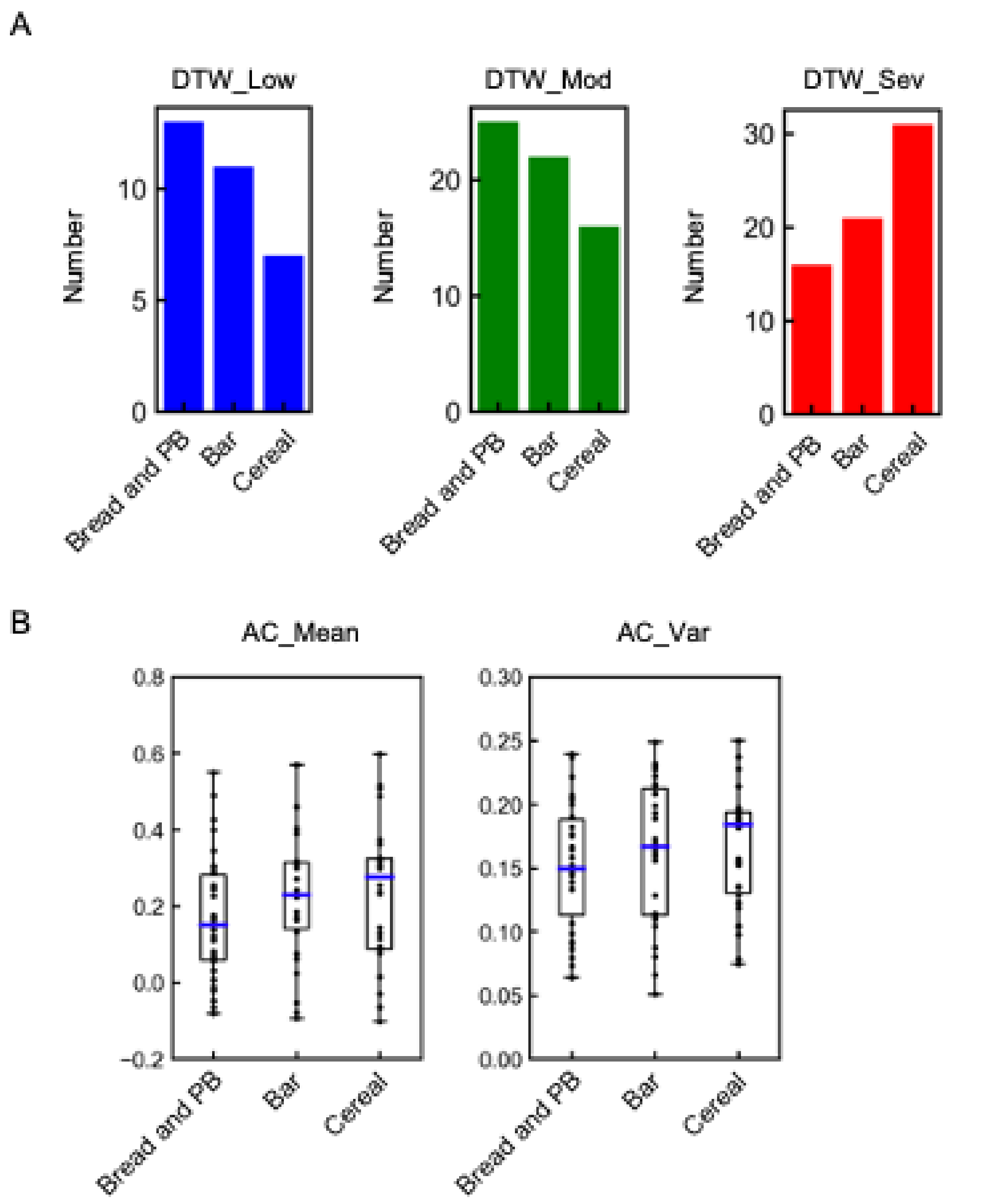

**Figure S12. Relation between AC_Mean, AC_Var, and standardized meals.**

A previously reported dataset[11] was used in this analysis. (A) Bar plots of DTW_Low (blue), DTW_Mod (green) and DTW_Sev (red) after eating the standardized meals. The values are derived from the previous study.[11] (B) Box plots of AC_Mean (the left) and AC_Var (the right) of each subject after eating the standardized meals. Each point corresponds to the value for a single subject, and blue lines indicate the median. Bread and PB, bread and peanut butter; Bar, PROBAR protein bar; Cereal, cornflakes and milk.

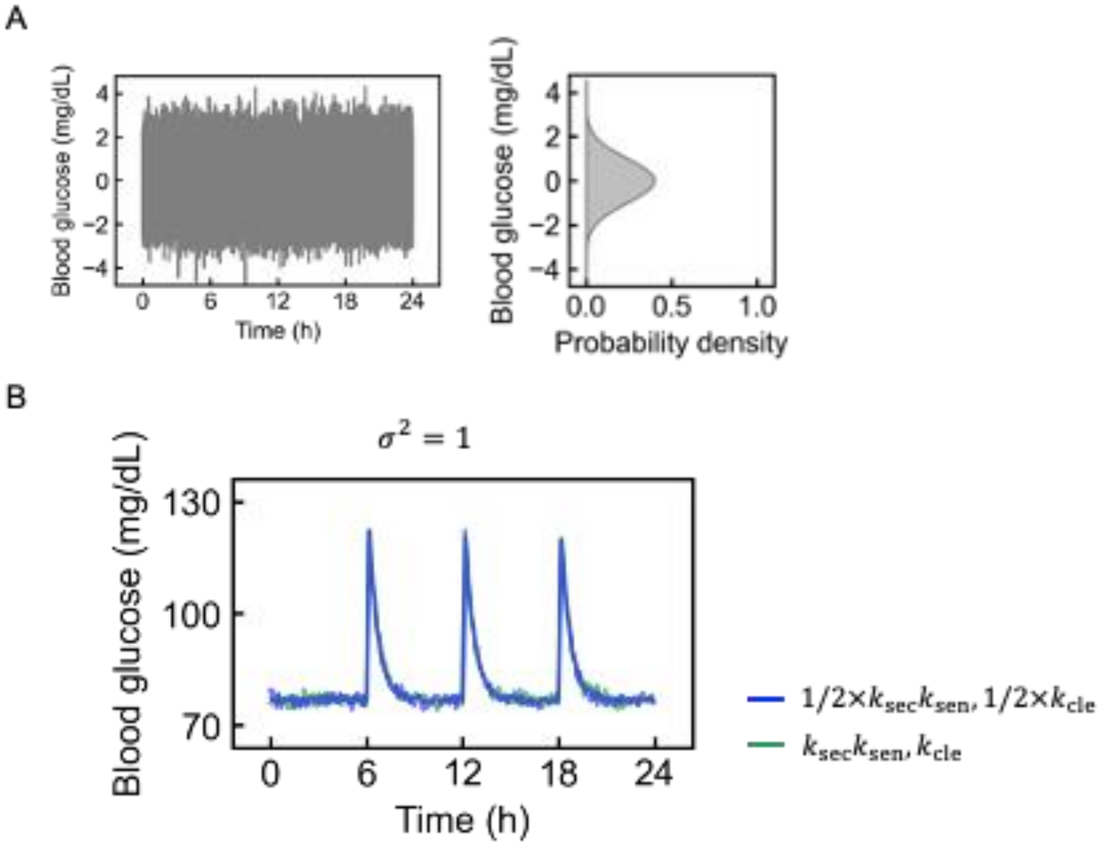

**Figure S13. Characterization of AC_Mean and AC_Var using simulated blood glucose with noise.**

(A) Zero-mean gaussian white noise with a variance of 1. The values of the noise added to the simulated glucose (the left) and the distribution of the noise (the right). (B) Twenty-four-hour simulated glucose concentration applied with gaussian white noise with the variances of 1. The colors of the lines are based on the values of $k_{\mathrm{sec}}k_{\mathrm{sen}}$ and $k_{\mathrm{cle}}$. The blue line is simulated using the parameters reported as the average values for healthy subjects.[12] The green line is simulated at half the values for healthy subjects.

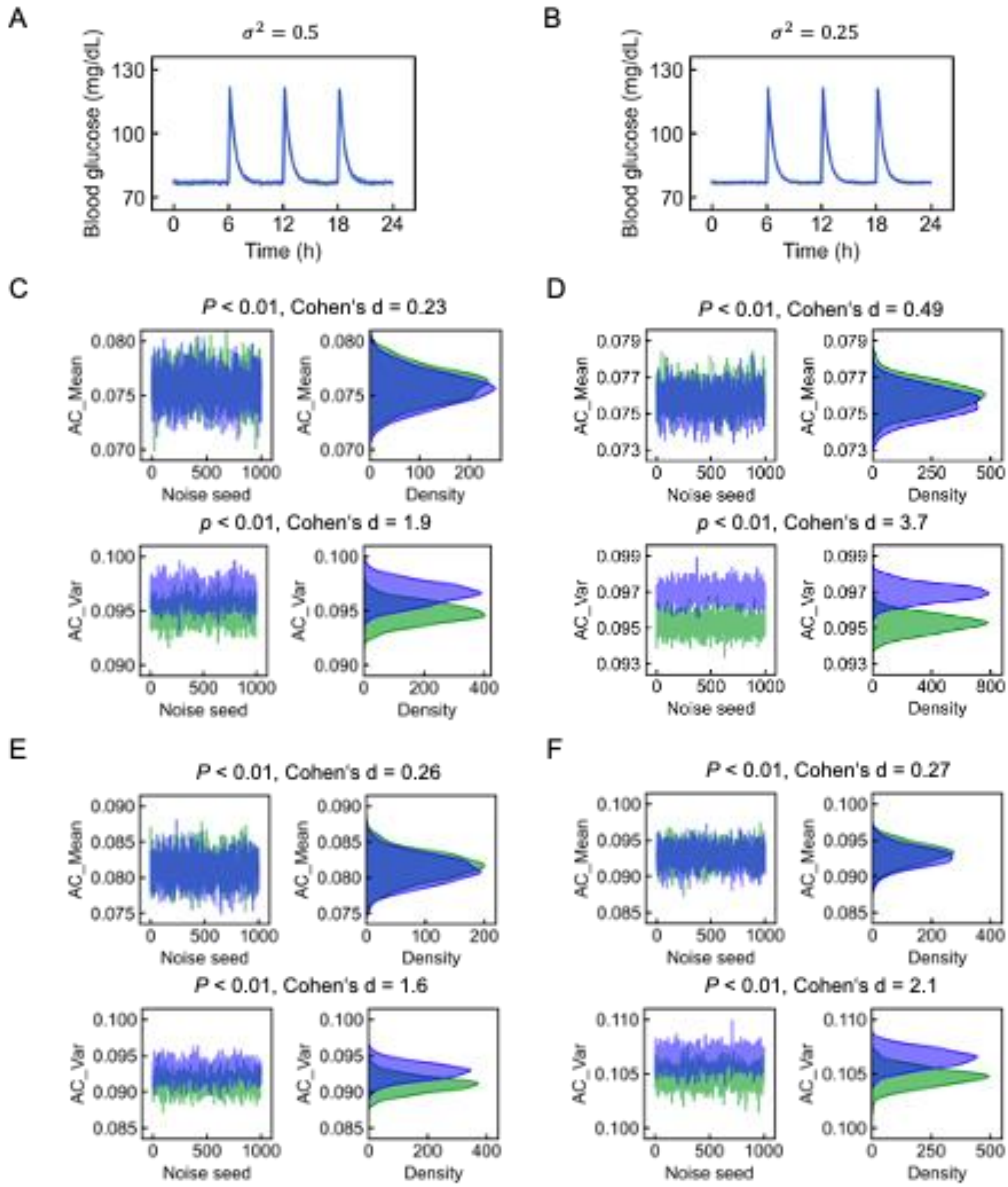

**Figure S14. Effects of noise and measurement conditions on AC_Mean and AC_Var.** (A, B) Twenty-four-hour simulated glucose concentration applied with gaussian white noise with the variances of 0.5 (A), and 0.25 (B). The colors of the lines are based on the values of $k_{\mathrm{sec}}k_{\mathrm{sen}}$ and $k_{\mathrm{cle}}$. The blue line is simulated using the parameters reported as the average values for healthy subjects.[12] The green line is simulated using half the values for healthy subjects.

(C, D) AC_Mean (the upper panels) and AC_Var (the lower panels) simulated from the 24-hour simulated glucose concentration with gaussian white noise with the variances of 0.5 (C) and 0.25 (D). The measurement interval was every 5 minutes. AC_Mean and AC_Var calculated in each trial are shown in the left panels. The distributions of AC_Mean and AC_Var are shown in the right. The *P* values are for testing the hypothesis of no difference between the two groups.

(E) AC_Mean (the upper panels) and AC_Var (the lower panels) simulated from the 72-hour simulated glucose concentration with gaussian white noise with the variance of 1. The measurement interval was every 5 minutes.

(F) AC_Mean (the upper panels) and AC_Var (the lower panels) simulated from the 24-hour simulated glucose concentration with gaussian white noise with the variance of 1. The measurement interval was every 1 minutes.

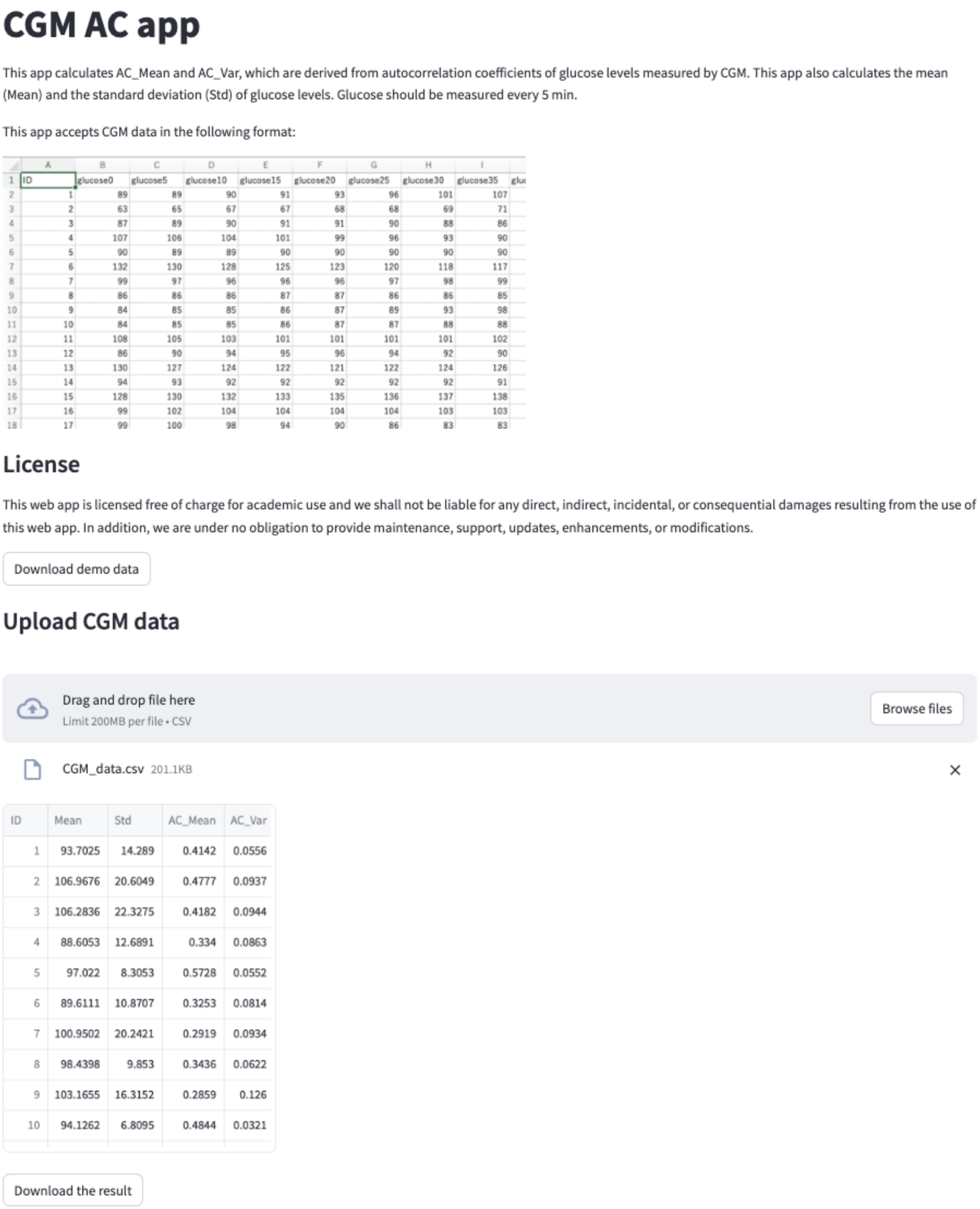

**Figure S15. Web application for calculating CGM-derived indices.**

This web application calculates AC_Mean and AC_Var. This application also calculates the mean (Mean) and the standard deviation (Std) of glucose levels. Glucose should be measured every 5 min. This application is implemented in streamlit.

#### Supplementary Tables

**Table S1. Characteristics of the subjects in this study.**

| | NGT | IGT | T2DM |
|---|---|---|---|
| Number of subjects (M/F) | 52 (38/14) | 9 (7/2) | 3 (3/0) |
| Age (years) | 35.3 ± 8.23 | 39.1 ± 13.6 | 43.3 ± 7.02 |
| BMI (kg/mm$^2$) | 22.3 ± 2.76 | 23.8 ± 3.21 | 26.3 ± 3.01 |
| Waist circumference (cm) | 78.7 ± 9.71 | 83.5 ± 9.48 | 90 ± 8.66 |
| Body fat percentage (%) | 21.8 ± 6.67 | 24 ± 4.84 | 23.5 ± 2.51 |
| SBP (mmHg) | 119 ± 15.8 | 132 ± 19.9 | 134 ± 27 |
| DBP (mmHg) | 79.2 ± 12.8 | 86.7 ± 13.5 | 94.8 ± 20.4 |
| TC (mg/dL) | 184 ± 29.4 | 223 ± 50.2 | 198 ± 44.3 |
| TG (mg/dL) | 82.9 ± 48.9 | 116 ± 66.9 | 166 ± 153 |
| LDL-C (mg/dL) | 110 ± 24.9 | 140 ± 40 | 118 ± 22.4 |
| HDL-C (mg/dL) | 57 ± 12.5 | 61 ± 23.3 | 50.7 ± 8.5 |
| FBG (mg/dL) | 91 ± 6.43 | 93.2 ± 7.95 | 116 ± 13.4 |
| HOMA-β | 111.7 ± 75.0 | 85.7 ± 44.5 | 101.7 ± 86.3 |
| HOMA-IR | 1.85 ± 0.99 | 1.64 ± 0.85 | 3.81 ± 2.90 |
| HbA1c (%) | 5.1 ± 0.234 | 5.31 ± 0.237 | 5.7 ± 0.529 |
| PG120 (mg/dL) | 110 ± 15.1 | 150 ± 11 | 223 ± 11 |
| I.I. | 0.969 ± 0.723 | 0.917 ± 0.911 | 0.657 ± 0.535 |
| Composite index | 5.86 ± 2.52 | 4.94 ± 2.6 | 4.34 ± 5.27 |
| Oral DI | 1.86 ± 0.605 | 1.77 ± 0.602 | 0.803 ± 0.211 |
| AUC_IRI | 353 ± 252 | 389 ± 333 | 284 ± 237 |
| ISI (x10$^{-4}$) | 12.9 ± 6.15 | 7.47 ± 3.62 | 8.97 ± 5.47 |
| Clamp DI | 40.8 ± 24.1 | 26.2 ± 21.5 | 13.9 ± 14.2 |
| $k_4$ (x10$^{-3}$) | 4.43 ± 8.51 | 3.30 ± 6.80 | 1.09 ± 1.29 |
| $k_5$ | 204 ± 1310 | 1030 ± 2270 | 163 ± 283 |
| $k_7$ | 306 ± 1480 | 244 ± 516 | 236 ± 408 |

Data are mean ± SD. NGT, normal glucose tolerance; IGT, impaired glucose tolerance; T2DM, type 2 diabetes mellitus; M, male; F, female; BMI, body mass index; SBP, systolic blood pressure; DBP, diastolic blood pressure; TC, total cholesterol; TG, triglycerides; LDL-C, low-density lipoprotein cholesterol; HDL-C, high-density lipoprotein cholesterol; FBG, fasting blood glucose; PG120, plasma glucose concentration at 120 min during the oral glucose tolerance test; I.I., insulinogenic index; oral DI, oral disposition index; AUC_IRI, area under insulin curve during the first 10 min of hyperglycemic clamp test; ISI, insulin sensitivity index; clamp DI; clamp disposition index. The indices $k_4$, $k_5$, and $k_7$ correspond to insulin sensitivity, insulin secretion, and insulin clearance, respectively.

### Table S2. List of significantly correlated relationships in Figure 4.

| Index1 | Index2 | Q |
|---|---|---|
| CONGA | CGM_Mean | 4.12E-31 |
| DTW_Sev | CGM_Mean | 4.12E-31 |
| ADRR | HBGI | 5.79E-31 |
| JINDEX | CGM_Mean | 3.21E-30 |
| MAGE | CGM_Std | 1.12E-29 |
| MAGE | LI | 2.98E-28 |
| DTW_Sev | JINDEX | 1.42E-27 |
| ADRR | JINDEX | 2.51E-25 |
| HBGI | CGM_Std | 8.07E-25 |
| DTW_Sev | GRADE | 1.02E-24 |
| ADRR | CGM_Std | 4.01E-24 |
| LI | CGM_Std | 1.28E-23 |
| GRADE | CGM_Mean | 1.11E-22 |
| MAG | LI | 2.05E-22 |
| ADRR | MAGE | 2.19E-21 |
| MAGE | HBGI | 2.93E-21 |
| DTW_Sev | DTW_Mod | 4.54E-21 |
| DTW_Sev | CONGA | 4.87E-21 |
| DTW_Mod | GRADE | 1.91E-20 |
| GRADE | JINDEX | 1.92E-20 |
| DBP | SBP | 5.94E-20 |
| MODD | CGM_Std | 5.95E-20 |
| Acir | BMI | 5.55E-19 |
| DTW_Low | MVALUE | 6.30E-19 |
| MODD | LI | 8.44E-19 |
| ADRR | LI | 8.78E-19 |
| GRADE | CONGA | 1.72E-18 |
| HBGI | LI | 2.01E-18 |
| HBGI | JINDEX | 5.81E-18 |
| LDL-C | TC | 1.04E-17 |
| JINDEX | CONGA | 1.36E-16 |
| MAGE | MODD | 5.46E-16 |
| DTW_Mod | JINDEX | 5.49E-15 |
| ADRR | MODD | 1.63E-14 |
| DTW_Sev | ADRR | 3.99E-14 |
| MAG | CGM_Std | 5.30E-14 |
| JINDEX | CGM_Std | 6.62E-14 |
| ADRR | CGM_Mean | 6.95E-14 |
| MODD | HBGI | 1.76E-13 |
| MAG | MAGE | 3.14E-13 |
| MVALUE | CONGA | 1.21E-12 |
| DTW_Low | CONGA | 1.39E-12 |
| DTW_Mod | CGM_Mean | 1.82E-12 |
| MAG | MODD | 3.00E-12 |
| MAG | ADRR | 1.37E-11 |
| AC_Var | AC_Mean | 2.66E-11 |
| MAGE | JINDEX | 3.64E-11 |
| MAG | HBGI | 5.80E-11 |
| JINDEX | LI | 1.12E-10 |
| ADRR | GRADE | 2.11E-10 |
| DTW_Mod | ADRR | 2.89E-10 |
| MODD | JINDEX | 4.01E-10 |
| k4 | ISI | 4.18E-10 |
| DTW_Low | CGM_Mean | 6.09E-10 |
| DTW_Sev | HBGI | 2.04E-09 |
| HBGI | CGM_Mean | 2.04E-09 |
| Clamp DI | AUC_IRI | 2.67E-09 |
| DTW_Mod | CGM_Std | 7.46E-09 |
| DTW_Mod | CONGA | 8.46E-09 |
| MVALUE | CGM_Mean | 1.09E-08 |
| AUC_IRI | I.I. | 1.79E-08 |
| Oral DI | FBG | 3.44E-08 |
| MAG | JINDEX | 3.90E-08 |
| DTW_Mod | HBGI | 6.51E-08 |
| DTW_Mod | MAGE | 1.82E-07 |
| DTW_Sev | CGM_Std | 2.15E-07 |
| ADRR | CONGA | 2.48E-07 |
| DTW_Mod | MODD | 2.78E-07 |
| DTW_Mod | LI | 2.81E-07 |
| k7 | Clamp DI | 3.29E-07 |
| DTW_Mod | MAG | 5.57E-07 |
| DTW_Sev | MVALUE | 5.87E-07 |
| DTW_Sev | DTW_Low | 6.38E-07 |
| GRADE | HBGI | 8.19E-07 |
| GRADE | CGM_Std | 8.30E-07 |
| DTW_Sev | MODD | 1.25E-06 |
| DTW_Sev | LI | 1.50E-06 |
| DTW_Sev | MAGE | 2.45E-06 |
| CGM_Std | CGM_Mean | 2.66E-06 |
| ISI | BMI | 6.29E-06 |
| k5 | AUC_IRI | 7.54E-06 |
| MODD | GRADE | 1.07E-05 |
| DTW_Sev | MAG | 1.19E-05 |
| MAGE | CGM_Mean | 2.35E-05 |
| LI | CGM_Mean | 2.62E-05 |
| DTW_Low | JINDEX | 4.07E-05 |
| Oral DI | I.I. | 4.36E-05 |
| MODD | CGM_Mean | 4.76E-05 |
| TG | BMI | 5.74E-05 |
| GRADE | LI | 6.21E-05 |
| HBGI | CONGA | 6.66E-05 |
| MAGE | GRADE | 6.93E-05 |
| MAG | GRADE | 7.21E-05 |
| MVALUE | GRADE | 7.25E-05 |
| Composite inde | BMI | 7.66E-05 |
| TG | Acir | 8.85E-05 |
| DTW_Low | GRADE | 0.000112486 |
| HbA1c | FBG | 0.000121215 |
| GRADE | HbA1c | 0.000142649 |
| Oral DI | HbA1c | 0.000142649 |
| k4 | BMI | 0.000247128 |
| ISI | Acir | 0.00024872 |
| DTW_Sev | HbA1c | 0.000253743 |
| Clamp DI | ISI | 0.00026014 |
| MAG | CGM_Mean | 0.00026014 |
| HDL-C | Acir | 0.000330646 |
| k7 | k5 | 0.000332808 |
| Composite inde | TG | 0.000478241 |
| k5 | I.I. | 0.000514828 |
| CGM_Mean | HbA1c | 0.000520933 |
| MVALUE | JINDEX | 0.000579571 |
| JINDEX | HbA1c | 0.000599176 |
| DTW_Sev | FBG | 0.000663652 |
| CONGA | HbA1c | 0.000665112 |
| TG | DBP | 0.000706576 |
| LDL-C | Acir | 0.000718272 |
| CGM_Mean | FBG | 0.00074586 |
| GRADE | FBG | 0.000753403 |
| k4 | Acir | 0.00077676 |
| DTW_Mod | HbA1c | 0.000992989 |
| ISI | Composite inde | 0.001085408 |
| Oral DI | HBGI | 0.001085408 |
| JINDEX | FBG | 0.001316391 |
| AUC_IRI | Composite inde | 0.001348712 |
| LDL-C | BMI | 0.00138484 |
| LDL-C | TG | 0.00142379 |
| k4 | TG | 0.001504461 |
| Oral DI | JINDEX | 0.001504461 |
| Clamp DI | I.I. | 0.001677858 |
| k7 | AUC_IRI | 0.001702246 |
| Oral DI | ADRR | 0.001729616 |
| I.I. | LI | 0.001945248 |
| AC_Var | LI | 0.002064181 |
| CONGA | FBG | 0.002064181 |
| ADRR | HbA1c | 0.002143157 |
| Composite inde | HDL-C | 0.002810709 |
| Clamp DI | TC | 0.002830998 |
| I.I. | MAGE | 0.003121567 |
| DTW_Mod | FBG | 0.003319991 |
| Oral DI | MAGE | 0.00339534 |
| AC_Var | MAGE | 0.00371404 |
| Oral DI | CGM_Std | 0.00371404 |
| PG120 | Acir | 0.003737898 |
| k4 | Composite inde | 0.003859299 |
| TG | TC | 0.004234987 |
| Oral DI | LI | 0.004336478 |
| k4 | DBP | 0.004411445 |
| TG | SBP | 0.004525636 |
| I.I. | ADRR | 0.004723324 |
| I.I. | HBGI | 0.005081873 |
| Oral DI | PG120 | 0.005286206 |
| BMI | SBP | 0.005867691 |
| Oral DI | CGM_Mean | 0.006026495 |
| HBGI | HbA1c | 0.006583487 |
| CONGA | CGM_Std | 0.006601782 |
| Oral DI | DTW_Sev | 0.007027917 |
| Composite inde | Acir | 0.007782074 |
| HDL-C | BMI | 0.007876389 |
| ISI | Body fat percer | 0.008033524 |
| k4 | Clamp DI | 0.008943411 |
| I.I. | CGM_Std | 0.009014746 |
| k7 | I.I. | 0.009014746 |
| I.I. | HbA1c | 0.009252925 |
| ADRR | FBG | 0.009405675 |
| PG120 | FBG | 0.009649787 |
| k7 | Acir | 0.009830415 |
| BMI | DBP | 0.010080663 |
| Oral DI | Acir | 0.010895023 |
| k4 | SBP | 0.012290776 |
| PG120 | CGM_Mean | 0.012472318 |
| DTW_Low | ADRR | 0.014767899 |
| PG120 | SBP | 0.015220923 |
| CGM_Std | HbA1c | 0.015464503 |
| k7 | Oral DI | 0.015469729 |
| Acir | SBP | 0.016939118 |
| Clamp DI | MAGE | 0.016939118 |
| HDL-C | TG | 0.016943615 |
| I.I. | MAG | 0.017013597 |
| PG120 | CONGA | 0.01725899 |
| Clamp DI | LI | 0.017845931 |
| ISI | TG | 0.017845931 |
| HbA1c | LDL-C | 0.017987234 |
| MODD | CONGA | 0.018836962 |
| PG120 | LDL-C | 0.019009105 |
| MODD | HbA1c | 0.019974258 |
| PG120 | HbA1c | 0.020220454 |
| PG120 | DTW_Sev | 0.021467789 |
| Oral DI | GRADE | 0.0216159 |
| PG120 | JINDEX | 0.0216159 |
| k5 | Clamp DI | 0.021751125 |
| PG120 | GRADE | 0.023756601 |
| PG120 | BMI | 0.02390429 |
| Composite inde | GRADE | 0.024276223 |
| Oral DI | MODD | 0.02427722 |
| LI | HbA1c | 0.024369851 |
| TC | BMI | 0.024380679 |
| Clamp DI | BMI | 0.024905418 |
| AC_Mean | Body fat percer | 0.025539657 |
| AUC_IRI | HDL-C | 0.026043434 |
| Oral DI | CONGA | 0.028585435 |
| Clamp DI | Oral DI | 0.029208947 |
| k7 | BMI | 0.030260159 |
| ISI | DBP | 0.030558647 |
| MAGE | CONGA | 0.031365965 |
| Composite inde | I.I. | 0.032145041 |
| MAGE | HbA1c | 0.032859464 |
| FBG | Acir | 0.033462832 |
| HBGI | FBG | 0.033595779 |
| Oral DI | BMI | 0.033595779 |
| Acir | DBP | 0.033838585 |
| CGM_Std | FBG | 0.035892037 |
| DTW_Mod | MVALUE | 0.035892037 |
| ISI | PG120 | 0.035892037 |
| Clamp DI | CGM_Std | 0.036845147 |
| ISI | SBP | 0.036845147 |
| PG120 | TC | 0.036845147 |
| Composite inde | CGM_Mean | 0.037860256 |
| CGM_Mean | Acir | 0.038022974 |
| I.I. | Body fat percer | 0.038022974 |
| k7 | AC_Var | 0.038022974 |
| LDL-C | SBP | 0.038022974 |
| Oral DI | DTW_Mod | 0.038022974 |
| Clamp DI | AC_Var | 0.038079506 |
| Composite inde | PG120 | 0.038079506 |
| Oral DI | TG | 0.038079506 |
| ISI | LDL-C | 0.038490701 |
| Oral DI | LDL-C | 0.038490701 |
| Clamp DI | LDL-C | 0.040097632 |
| k4 | AC_Var | 0.040932358 |
| AUC_IRI | MAGE | 0.041223807 |
| Composite inde | SBP | 0.04245559 |
| I.I. | MODD | 0.042994604 |
| k7 | ISI | 0.043499242 |
| Oral DI | MAG | 0.044698865 |
| DTW_Sev | Acir | 0.044977901 |
| CONGA | Acir | 0.045649698 |
| LDL-C | DBP | 0.045649698 |
| TC | DBP | 0.045649698 |
| MAG | FBG | 0.045824416 |
| Clamp DI | ADRR | 0.046236045 |
| LI | CONGA | 0.046681438 |
| k7 | LDL-C | 0.047155766 |
| FBG | BMI | 0.047826039 |
| k4 | LDL-C | 0.048417599 |
| Clamp DI | MODD | 0.048654305 |
| TC | SBP | 0.04865957 |

### Table S3. List of significantly correlated relationships in Figure 5E.

| Index1 | Index2 | Q |
|---|---|---|
| CONGA | CGM_Mean | 2.33E-23 |
| CGM_Mean | DTW_Sev | 4.92E-22 |
| JINDEX | CGM_Mean | 4.92E-22 |
| JINDEX | DTW_Sev | 8.42E-21 |
| CGM_Mean | DTW_Low | 5.36E-19 |
| CONGA | DTW_Low | 1.58E-18 |
| CONGA | DTW_Sev | 4.72E-15 |
| ADRR | HBGI | 5.84E-15 |
| JINDEX | CONGA | 2.67E-14 |
| MVALUE | DTW_Low | 5.20E-14 |
| LI | CGM_Std | 2.68E-13 |
| DTW_Sev | DTW_Low | 3.06E-13 |
| MODD | CGM_Std | 6.76E-13 |
| LDL-C | TC | 7.77E-13 |
| ADRR | CGM_Std | 2.16E-12 |
| HBGI | CGM_Std | 2.29E-12 |
| MVALUE | CONGA | 2.50E-12 |
| GRADE | DTW_Mod | 3.02E-12 |
| ADRR | JINDEX | 3.23E-12 |
| ADRR | LI | 3.72E-12 |
| JINDEX | DTW_Low | 1.62E-11 |
| MAGE | LI | 4.70E-11 |
| MAGE | CGM_Std | 5.29E-11 |
| MVALUE | CGM_Mean | 2.17E-10 |
| AC_Var | AC_Mean | 1.11E-09 |
| MODD | LI | 8.07E-09 |
| GRADE | DTW_Sev | 8.67E-09 |
| ADRR | MAGE | 1.77E-08 |
| ADRR | DTW_Sev | 2.01E-08 |
| ADRR | CGM_Mean | 2.23E-08 |
| HBGI | LI | 2.34E-08 |
| MVALUE | DTW_Sev | 2.88E-08 |
| ADRR | MODD | 7.94E-08 |
| AC_Var | LI | 7.94E-08 |
| MAGE | MODD | 1.29E-07 |
| JINDEX | CGM_Std | 1.41E-07 |
| MAGE | HBGI | 1.53E-07 |
| HBGI | JINDEX | 2.14E-07 |
| GRADE | JINDEX | 4.83E-07 |
| JINDEX | LI | 8.71E-07 |
| AC_Mean | LI | 1.10E-06 |
| AC_Var | ADRR | 1.81E-06 |
| GRADE | CGM_Mean | 1.99E-06 |
| MVALUE | JINDEX | 2.02E-06 |
| MODD | JINDEX | 2.42E-06 |
| PG120 | DTW_Low | 3.85E-06 |
| PG120 | CGM_Mean | 3.87E-06 |
| MODD | HBGI | 7.98E-06 |
| AC_Var | HBGI | 7.98E-06 |
| LI | DTW_Sev | 2.01E-05 |
| PG120 | JINDEX | 2.01E-05 |
| ADRR | DTW_Low | 2.03E-05 |
| ADRR | CONGA | 2.40E-05 |
| PG120 | CONGA | 2.70E-05 |
| GRADE | CONGA | 2.93E-05 |
| CGM_Std | DTW_Sev | 3.38E-05 |
| PG120 | DTW_Sev | 4.24E-05 |
| HBGI | CGM_Mean | 6.23E-05 |
| GRADE | CGM_Std | 7.23E-05 |
| PG120 | ADRR | 7.94E-05 |
| PG120 | AC_Var | 9.86E-05 |
| PG120 | MVALUE | 9.98E-05 |
| CGM_Mean | BMI | 0.000103122 |
| HBGI | DTW_Sev | 0.000149063 |
| AC_Var | MAGE | 0.000161274 |
| DTW_Low | BMI | 0.000177644 |
| LI | CGM_Mean | 0.000178295 |
| MODD | DTW_Sev | 0.000178295 |
| CGM_Std | CGM_Mean | 0.000209523 |
| DTW_Sev | BMI | 0.000216105 |
| CONGA | BMI | 0.000252547 |
| MAG | LI | 0.000288568 |
| GRADE | DTW_Low | 0.000320626 |
| MAGE | JINDEX | 0.000346641 |
| AC_Var | CGM_Std | 0.000361397 |
| ADRR | GRADE | 0.000361535 |
| SSPG | HDL-C | 0.000361535 |
| MVALUE | BMI | 0.00036185 |
| SSPG | BMI | 0.000385325 |
| MAG | CGM_Std | 0.000408763 |
| JINDEX | BMI | 0.000557693 |
| DTW_Sev | DTW_Mod | 0.000719491 |
| MODD | CGM_Mean | 0.000764515 |
| PG120 | HbA1c | 0.00087973 |
| MAG | GRADE | 0.000986011 |
| AC_Mean | ADRR | 0.001236288 |
| PG120 | LI | 0.0012932 |
| MODD | GRADE | 0.001711185 |
| PG120 | HBGI | 0.001966987 |
| PG120 | AC_Mean | 0.002162977 |
| GRADE | LI | 0.002268089 |
| HBGI | CONGA | 0.002308231 |
| AC_Mean | MAGE | 0.002376012 |
| GRADE | HBGI | 0.002401954 |
| MAG | DTW_Sev | 0.002401954 |
| CONGA | HbA1c | 0.002587043 |
| CGM_Std | DTW_Mod | 0.002617752 |
| FBG | HbA1c | 0.002789005 |
| JINDEX | DTW_Mod | 0.002789005 |
| PG120 | BMI | 0.003272172 |
| SSPG | DTW_Low | 0.003379039 |
| MAG | MODD | 0.003494856 |
| PG120 | SSPG | 0.003892529 |
| AC_Var | JINDEX | 0.004072663 |
| CGM_Mean | HbA1c | 0.004115984 |
| SSPG | MVALUE | 0.004131666 |
| HBGI | DTW_Low | 0.004383312 |
| SSPG | AC_Var | 0.004383312 |
| MAG | ADRR | 0.004728298 |
| MAGE | DTW_Sev | 0.00537048 |
| SSPG | CGM_Mean | 0.00537048 |
| MAG | JINDEX | 0.0063552 |
| AC_Var | HbA1c | 0.0063552 |
| SSPG | HBGI | 0.0063552 |
| MVALUE | HbA1c | 0.006799826 |
| SSPG | CONGA | 0.007175 |
| GRADE | BMI | 0.007899975 |
| MAG | MAGE | 0.008099887 |
| PG120 | FBG | 0.008202642 |
| MVALUE | ADRR | 0.008304901 |
| CONGA | CGM_Std | 0.008305199 |
| AC_Var | CGM_Mean | 0.00855102 |
| DTW_Low | HbA1c | 0.008642156 |
| PG120 | CGM_Std | 0.009161506 |
| LI | DTW_Low | 0.009621334 |
| CGM_Mean | DTW_Mod | 0.010175349 |
| MAGE | CGM_Mean | 0.010175349 |
| SSPG | LDL-C | 0.010239365 |
| CONGA | FBG | 0.010322657 |
| MODD | CONGA | 0.0109737 |
| AC_Var | DTW_Sev | 0.012097202 |
| HDL-C | TG | 0.012634482 |
| AC_Var | MODD | 0.012848983 |
| SSPG | ADRR | 0.012848983 |
| CGM_Mean | FBG | 0.013923283 |
| LI | CONGA | 0.014276773 |
| HBGI | DTW_Mod | 0.014276773 |
| JINDEX | HbA1c | 0.014481248 |
| AC_Var | FBG | 0.015222803 |
| SSPG | JINDEX | 0.016370604 |
| DTW_Sev | FBG | 0.018050087 |
| AC_Mean | HBGI | 0.018476648 |
| PG120 | MAGE | 0.018476648 |
| LI | BMI | 0.019367416 |
| AC_Mean | CGM_Std | 0.019367416 |
| JINDEX | FBG | 0.01989891 |
| MVALUE | GRADE | 0.020887155 |
| DTW_Sev | HbA1c | 0.02089033 |
| LI | DTW_Mod | 0.021153456 |
| MAGE | GRADE | 0.021593064 |
| ADRR | DTW_Mod | 0.021593064 |
| ADRR | BMI | 0.02175833 |
| TG | HbA1c | 0.02198786 |
| CGM_Std | DTW_Low | 0.02486039 |
| AC_Mean | BMI | 0.02486039 |
| AC_Mean | MAG | 0.026204208 |
| MAG | CGM_Mean | 0.027693916 |
| TG | TC | 0.028910927 |
| AC_Var | TG | 0.030760645 |
| HDL-C | BMI | 0.033023807 |
| CONGA | DTW_Mod | 0.03413692 |
| SSPG | DTW_Sev | 0.034544995 |
| AC_Var | DTW_Low | 0.035165679 |
| MAG | DTW_Mod | 0.037564835 |
| AC_Mean | DTW_Sev | 0.038721889 |
| PG120 | MODD | 0.038721889 |
| DTW_Low | FBG | 0.040507648 |
| MAG | HBGI | 0.040507648 |
| SSPG | AC_Mean | 0.041805569 |
| AC_Mean | JINDEX | 0.04345714 |
| MAG | DTW_Low | 0.043787903 |
| MODD | DTW_Mod | 0.04642888 |
| SSPG | LI | 0.047075889 |
| MODD | DTW_Low | 0.048117514 |
| CGM_Std | BMI | 0.048206535 |
| MODD | TC | 0.048206535 |
| MVALUE | FBG | 0.048206535 |
| HBGI | BMI | 0.048303972 |
